# Association of GLP-1 Receptor Agonist Prescriptions and Alcohol Consumption in the National Institutes of Health’s *All of Us* Cohort

**DOI:** 10.64898/2026.01.15.26344218

**Authors:** Benjamin Tyndall, Angela Gasdaska, M. Daniel Brannock, Ed Preble, Melissa McPheeters, Laura Marcial, Ariba Huda, Josephine Egan, Tamara R. Litwin, Jennifer Adjemian, Chandan Sastry, Mehdi Farokhnia, Lorenzo Leggio

**Affiliations:** RTI International, Durham, NC; National Institutes of Health, Bethesda, MD

## Abstract

**Importance:** Alcohol use is a leading cause of morbidity and mortality worldwide. Growing evidence suggests that glucagon-like peptide-1 receptor agonists (GLP-1RAs) may represent a novel potential pharmacotherapeutic tool for alcohol use disorder (AUD).

**Objective:** To examine the association between GLP-1RA prescriptions and alcohol use.

**Design:** This cohort study used a cross-sectional measure of alcohol consumption and longitudinal electronic health record (EHR) data collected between 1981 and October 2023 from NIH’s *All of Us* Research Program participants.

**Setting:** *All of Us* is a large program to recruit and collect surveys, EHR, genomic, and wearable data from a wide array of Americans. The data presented here are from the *All of Us* Curated Data Repository version 8.

**Participants:** 393,596 *All of Us* participants with EHR data recruited across the United States.

**Exposure:** At least two GLP-1RA prescription records in the EHR.

**Main Outcomes and Measures:** Alcohol Use Disorders Identification Test (AUDIT-C) scores and responses to individual AUDIT-C questions.

**Results:** Among 15,447 participants with at least two recorded GLP-1RA prescriptions on separate days, 3650 had active GLP-1RA prescriptions, 5642 would have future GLP-1RA prescriptions (primary comparison group), and 544 had former GLP-1RA prescriptions. Those with active GLP-1RA prescriptions had statistically significant but modestly lower AUDIT-C scores on average compared with those with future prescriptions (incidence rate ratio [IRR] of 0.95; 95% CI, 0.91-0.99; *P* = 0.01). Participants with a former GLP-1RA prescription had lower AUDIT-C scores compared with those with future prescriptions, but this difference was not statistically significant. Results were similar using a propensity-score matched comparison group with a lower average AUDIT-C score for the current GLP-1RA group (IRR = 0.89; 95% CI, 0.85-0.93; *P* = <0.001) and no significant difference for the former prescription group. Analysis of individual AUDIT-C questions shows a significant association with GLP-1RA prescriptions and frequency of drinking but not drinks per occasion or binge drinking.

**Conclusions and Relevance:** This study’s findings indicate that GLP-1RAs may reduce alcohol consumption by decreasing use frequency. Experimental studies and randomized controlled trials are needed to test the mechanisms and potential efficacy of GLP-1RAs in people with AUD.

**KEY POINTS:** *Question:* Is there an association between glucagon-like peptide-1 receptor agonist (GLP-1RA) prescriptions and alcohol consumption?

*Findings:* In this observational cohort study of 15 447 people with GLP-1RA prescriptions in NIH’s *All of Us* cohort, active GLP-1RA prescriptions were associated with significantly lower Alcohol Use Disorders Identification Test (AUDIT-C) scores compared with scores of those who would have GLP-1RAs in the future and of a matched comparison group with no GLP-1RAs. People with former GLP-1RAs did not have lower AUDIT-C scores than these comparison groups.

*Meaning:* Active GLP-1RA use may be effective in reducing alcohol consumption.

## Introduction

Glucagon-like peptide-1 receptor agonists (GLP-1RAs) have been approved by the U.S. Food and Drug Administration (FDA) to treat type 2 diabetes, weight loss, and obstructive sleep apnea. GLP-1RAs are being studied as potential pharmacotherapies for other conditions including alcohol use disorder (AUD) and other substance use disorders.^1^ Alcohol is one of the most widely used addictive substances, and alcohol misuse contributes to almost 178,000 deaths annually in the United States, making it a leading cause of mortality and morbidity.^2–4^ Yet, only three pharmacotherapies are approved for AUD, they are not widely used, and not all patients with AUD respond to them; therefore, discovering additional pharmacotherapies for AUD is paramount.^5^

The mechanisms through which GLP-1RAs could reduce alcohol consumption are incompletely understood but may relate to GLP-1RA’s effects on appetite/satiety, reward processing, stress regulation, cognition, or neuroinflammation.^1,6^ Animal studies show that GLP- 1RA administration leads to reduction in alcohol consumption and other alcohol-related outcomes.^1,7^ Early observational studies using electronic health record (EHR) data support GLP- 1RAs’ potential to reduce alcohol consumption in humans as well as lower risk of an AUD diagnosis and alcohol-related events such as intoxication and hospitalizations.^8–12^ However, most EHR-based studies lack consistent alcohol use measures and frequencies, limiting the ability to quantitatively study alcohol use and investigate associations where alcohol use is not clinically observed but still relevant to health outcomes. One study found that GLP-1RAs were associated with lowered alcohol consumption among a mostly male cohort of veterans using EHRs from the U.S. Veterans Health Administration, whereas another found decreased use in a largely female cohort enrolled in a single weight-loss program.^13,14^ Findings about alcohol use among more diverse patient populations are limited.

In this study, we used data from NIH’s *All of Us* Research Program to address gaps in understanding the association between GLP-1RAs and self-reported alcohol consumption. *All of Us* includes a longitudinal health database that has enrolled over 633,000 people living in the United States and its territories and collects data from surveys, EHRs, physical measurements, wearables, and biospecimens.^15^ Surveys from *All of Us* include questions about alcohol consumption, and linked EHR data provide longitudinal health histories for most participants, including drug exposures, laboratory tests, and diagnoses. We used this data resource to examine self-reported alcohol use and GLP-1RA initiation within a broad population to identify whether, and to what extent, GLP-1RAs are associated with alcohol consumption.

## METHODS

### Data Source

This study used Controlled Tier data from the *All of Us* Curated Data Repository version 8 (CDRv8).^16^ CDRv8 data contain information on 633,547 participants who reside in the United States, were 18 or older at the time of enrollment in *All of Us*, and enrolled between May 6, 2018, and October 1, 2023. All participant data were collected between 1981 and October 1, 2023, with histories for most participants beginning between 2014 and 2019.

For this study, we used surveys, EHR, and physical measurement data. The baseline surveys include sociodemographics, health, and lifestyle questions. CDRv8 includes dated survey responses for 633,547 participants, including 578,451 participants who answered the Lifestyle survey between May 31, 2017, and September 30, 2023.^17^ To date, participants have provided only one response per survey type while enrolled in the program.^16^ EHR data in *All of Us* are available for over 393,000 participants. The EHR data are prospectively and retrospectively longitudinal, with some participants providing records dating to 1980. *All of Us* has also collected physical measurements, such as height, weight, body mass index (BMI), heart rate, blood pressure, and hip and waist circumference for over 509,000 participants.

### Study Population

The study population included participants who responded to the Lifestyle survey; had a documented or calculable BMI; had at least one inpatient, outpatient, or emergency department (ED) visit in the 2 years before survey completion (see eTable 1 for visit type definitions); and had no history of medullary thyroid carcinoma, multiple endocrine neoplasia syndrome type 2, or end-stage renal disease by the time of survey completion. We excluded participants with these conditions because they are contraindications to GLP-1RA use.^18,19^ We also excluded participants with any recorded prescription of the three FDA-approved medications to treat AUD (naltrexone, acamprosate, and disulfiram)^20^; those with any record of pregnancy in the year before survey completion; and those who underwent bariatric surgery because potential changes in alcohol consumption in these groups could not be distinguished from the potential effects of GLP-1RAs. Finally, we excluded participants with self-reported zero lifetime alcohol consumption.

### Exposure Groups

We defined two exposed groups: those with an active GLP-1RA prescription (active prescription group) and those with a recent but former GLP-1RA prescription (former prescription group). Exposed groups had at least two GLP-1RA prescriptions on separate days with at least one record within the 365 days preceding Lifestyle survey completion. GLP-1RAs were identified as any drugs containing albiglutide, dulaglutide, exenatide, liraglutide, lixisenatide, semaglutide, and the GLP-1/gastric inhibitory polypeptide (GIP) dual agonist tirzepatide. (eTable 2 contains the Observational Medical Outcomes Partnership [OMOP] concept IDs used to identify these medications in the *All of Us* drug exposure table.) For this analysis, we used all records in the drug exposure table, including prescriptions written by a clinician, dispensations, claims, administrations, medication list entries, self-reports, and those without a specified source, and refer to them collectively as “prescriptions.” In the active prescription group, the first GLP-1RA record did not occur in the 1 month before survey completion, and the last GLP-1RA record was no more than 90 days before Lifestyle survey completion or occurred after survey completion. In the former prescription group, the last GLP- 1RA record occurred between 90 and 365 days before taking the Lifestyle survey.

We defined two unexposed groups based on an absence of GLP-1RA records before survey completion. Our primary unexposed group, the future prescription group, consisted of participants with no GLP-1RA records before completing the Lifestyle survey but with at least two GLP-1RA prescriptions on separate days concurrent with or after completing the survey. This unexposed comparison group replaces a traditional baseline comparison, as longitudinal survey results are not available in this dataset. This group is substantially more similar to participants in the exposed groups than participants with no lifetime GLP-1RA records. We defined a secondary unexposed group that consists of anyone in the aforementioned study population with no GLP-1RA record exposures at the time of survey completion (not excluding those with future GLP-1RA exposures and no requirement for this) matched to our exposed groups on our full set of covariates described later using propensity-score matching (see Figure 1).

**Figure 1.**
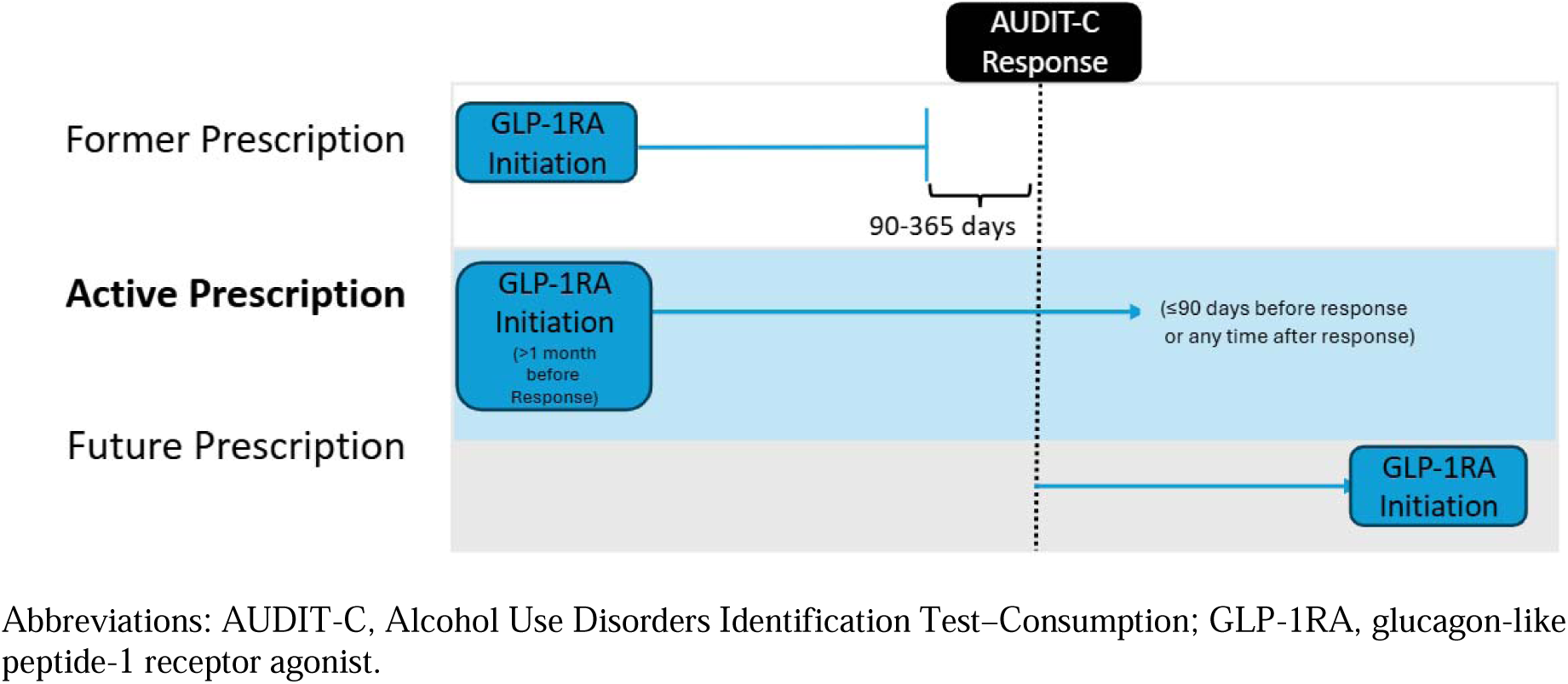
Exposure and Comparison Groups. Abbreviations: AUDIT-C, Alcohol Use Disorders Identification Test–Consumption; GLP-1RA, glucagon-like peptide-1 receptor agonist.

### Outcome

Our primary outcome was Alcohol Use Disorders Identification Test–Consumption (AUDIT-C) score. The AUDIT-C is an alcohol screening tool used to screen for and assess alcohol use.^21^ It includes three questions, each scored from 0 to 4; the total score ranges from 0 to 12. Scores denote severity of alcohol use: 0 indicates no use, 1–3 low-risk drinking, 4–7 at-risk drinking, 8-12 hazardous or heavy episodic use.

We derived AUDIT-C scores using three questions from the Lifestyle survey:

1. *How often did you have a drink containing alcohol in the past year?*
2. *On a typical day when you drink, how many drinks do you have?*
3. *How often did you have six or more drinks on one occasion in the past year?*

These questions were asked of all participants, regardless of sex and age. Each participant answered the survey only once.

### Covariates

We defined covariates using survey, EHR, and physical measurement data. We included covariates for sex, race, ethnicity, education, household income, and health insurance type from the Basics survey. We included any history of substance use (including tobacco) from the Lifestyle survey. We calculated participant age at the time of the Lifestyle survey using date of birth. We included BMI calculated or recorded closest to the Lifestyle survey.

Using EHR data, we defined covariates for history of type 1 diabetes, type 2 diabetes, depression, or liver disease, as any individual who had at least one diagnosis concurrent with or before Lifestyle survey completion (OMOP concept IDs used to define these conditions are listed in eTable 1). We defined hypertension as at least one hypertension diagnosis or at least two hypertensive systolic or diastolic blood pressure measurements before or concurrent with the Lifestyle survey. We controlled for comorbidities observed any time before the Lifestyle survey using a modified Charlson Comorbidity Index, excluding liver disease and diabetes using the ICD-9 and ICD-10 codes (see eTable 3, 4, 5^23^). Additionally, we controlled for any record of on- or off-label obesity treatments or non-FDA-approved off-label AUD treatments in the year preceding the Lifestyle survey (see eTable 1). Finally, we derived measures of health care utilization by calculating the total number of inpatient, outpatient, or ED visits in the 2 years preceding the Lifestyle survey.

We excluded individuals missing a BMI measurement or responses to the Lifestyle survey questions necessary to create our outcome measure. For all other covariates based on survey questions, individuals missing data were included in the “skipped” or “prefer not to answer” categories.

### Statistical Analysis

The primary outcome was the incident rate ratio (IRR) of GLP-1RA exposure on AUDIT-C scores. The IRR was calculated from two weighted, multivariate, negative binomial models that compared each of the two exposed groups (active and former) with the future prescription group described earlier. Each model used the AUDIT-C score as the dependent variable, the covariates described earlier as independent variables, and weights defined from propensity of exposure models. The propensity models were logistic regressions that used the same set of covariates as independent variables and exposure status as the dependent variable. Individuals were assigned weights corresponding to their inverse probabilities of being in their respective exposed group.

We conducted three secondary analyses. The first analysis considered each of the three composite AUDIT-C questions separately as outcomes. Each effect was calculated alone in the primary analysis. The second secondary analysis compared the exposed groups with the matched unexposed comparison group, using one-to-one propensity-score matching and the same set of covariates as the primary analysis. The third secondary analysis replicated the primary analysis without incorporating propensity weights. Analyses were conducted using a Jupyter Notebook workspace, Python version 3.10.16, and R version 4.5.0 in the *All of Us* Researcher Workbench.

## Results

Of the 393,596 *All of Us* participants with EHR data available, we identified 20,768 who had at least one GLP-1RA record. Of these, 15,447 had at least two GLP-1RA records on separate days. After applying all exclusion criteria and grouping by dates of GLP-1RA records in relation to Lifestyle survey completion date, we had 3650 in our active GLP-1RA prescription, 544 in our former GLP-1RA prescription, and 5642 in our future GLP-1RA prescription (primary unexposed group) groups (Figure 2). 270,324 All of Us participants met the inclusion criteria to be considered for our matched comparison group (eFigure 1).

**Figure 2.**
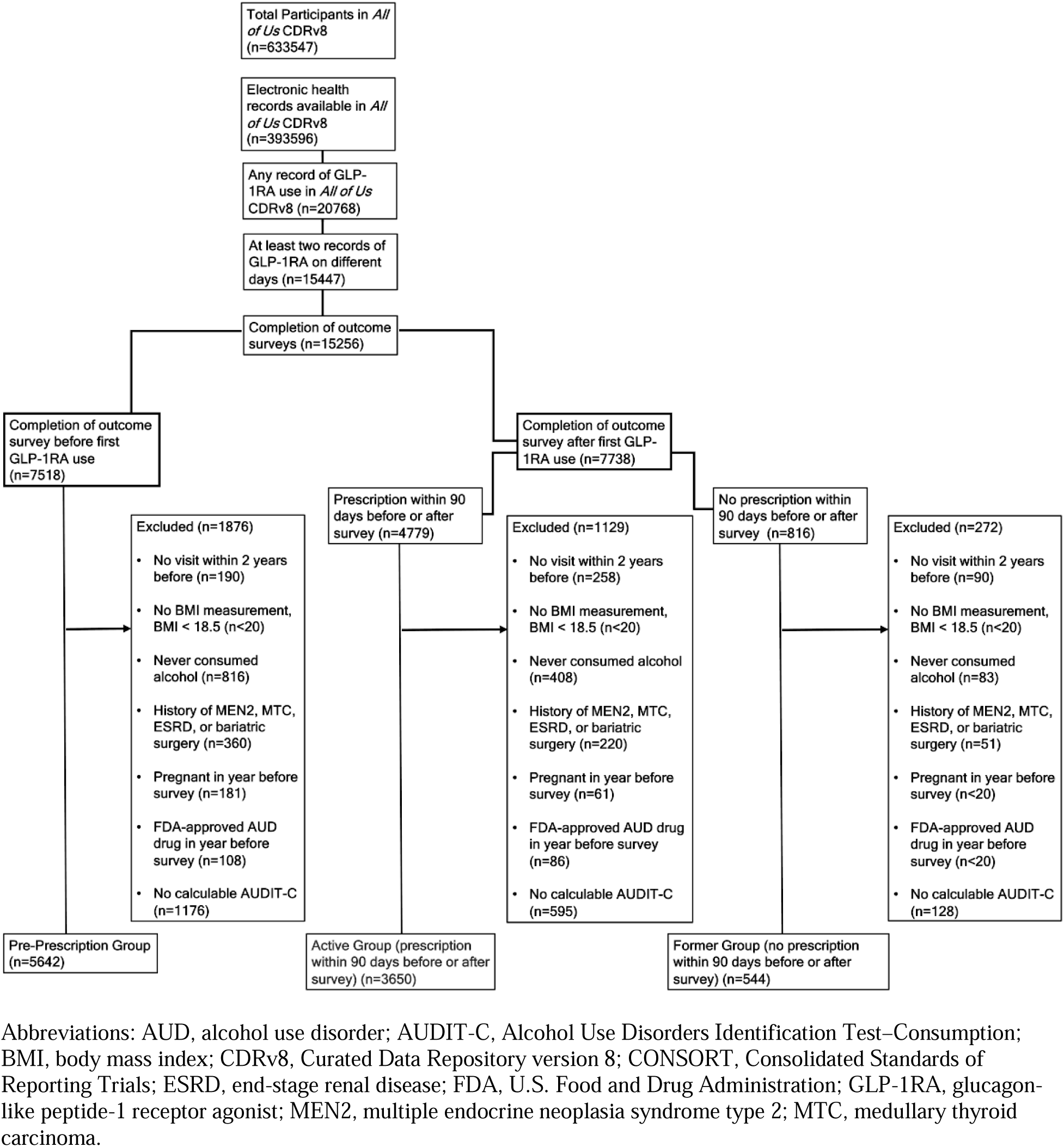
**CONSORT Diagram Showing Attrition from Exclusion Criteria** Abbreviations: AUD, alcohol use disorder; AUDIT-C, Alcohol Use Disorders Identification Test–Consumption; BMI, body mass index; CDRv8, Curated Data Repository version 8; CONSORT, Consolidated Standards of Reporting Trials; ESRD, end-stage renal disease; FDA, U.S. Food and Drug Administration; GLP-1RA, glucagon-like peptide-1 receptor agonist; MEN2, multiple endocrine neoplasia syndrome type 2; MTC, medullary thyroid carcinoma.

Characteristics differed between our active and former exposed groups and our unexposed future prescription group (Table 1). After propensity-score weighting separately for each exposed group, groups were well balanced, having absolute standardized mean differences (SMDs) of less than 0.1 for all characteristics (eFigures 2 and 3). For our secondary analyses, we performed propensity-score matching. After propensity-score matching, our matched unexposed groups were well balanced, with SMDs of less than 0.1 for all characteristics but former tobacco use (eFigures 4 and 5).

**Figure 3.**
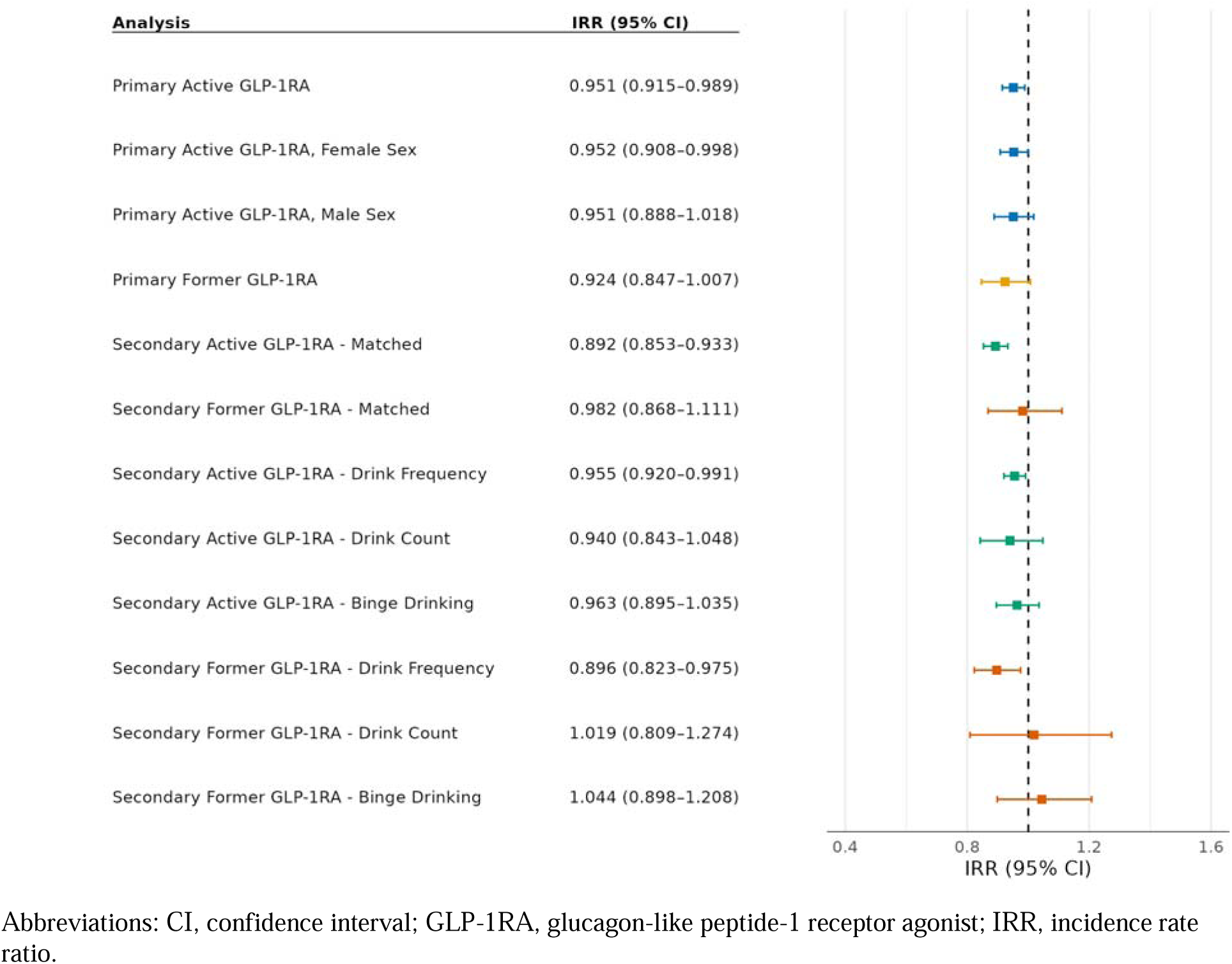
**Forest Plot of Key Effect Estimates** Abbreviations: CI, confidence interval; GLP-1RA, glucagon-like peptide-1 receptor agonist; IRR, incidence rate ratio.

**Table 1.**
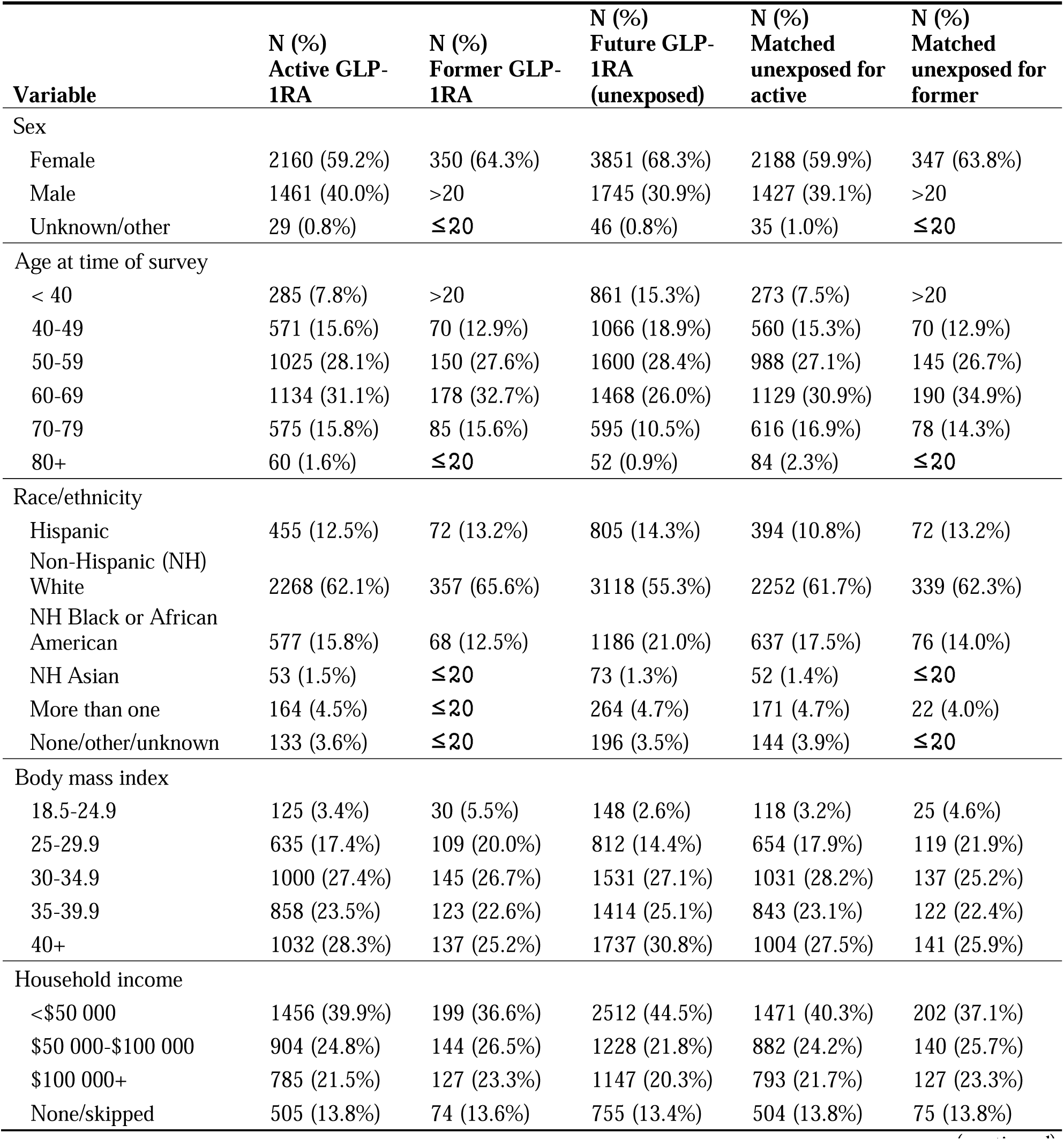
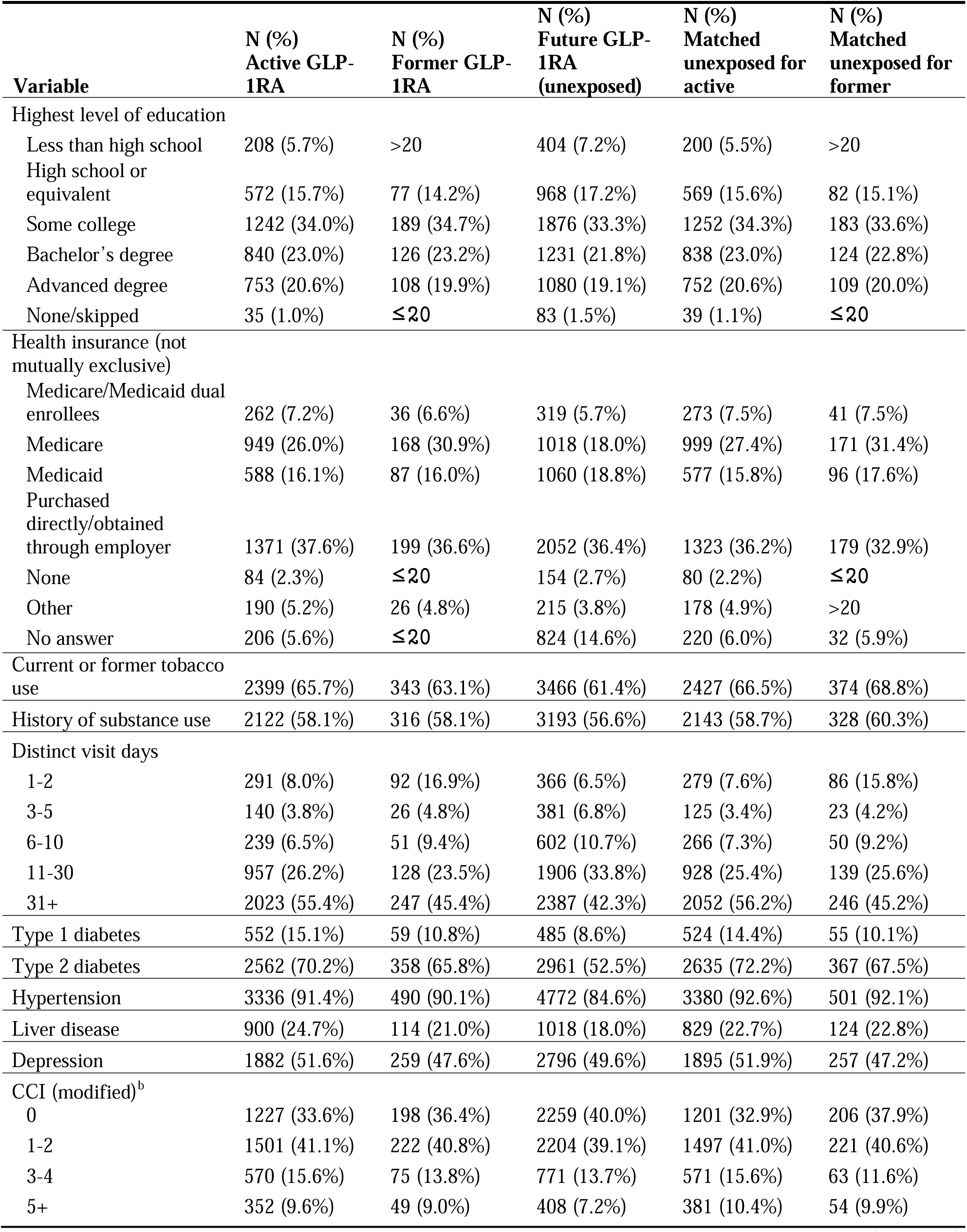
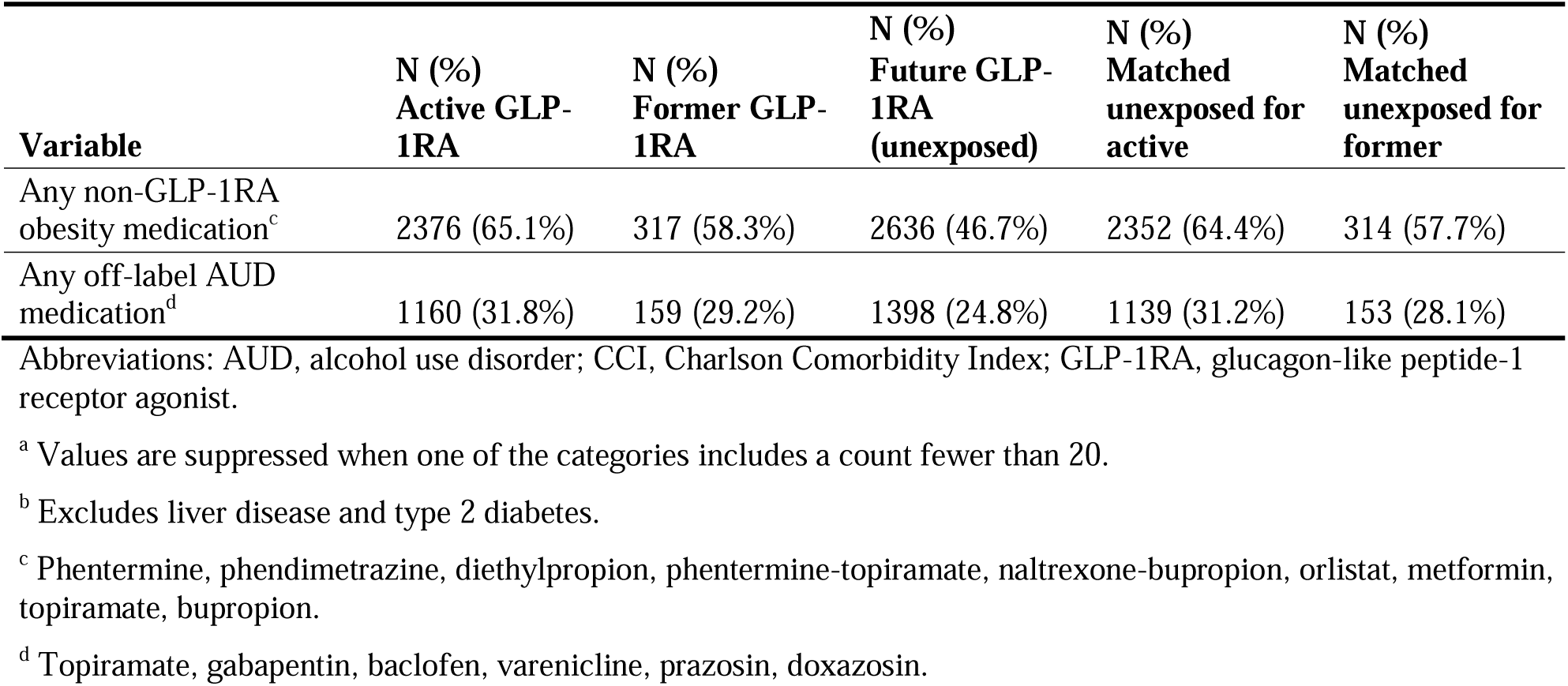
Cohort Characteristics for Each Exposed Group and Unexposed Comparison Groups^a^.

We used weighted negative binomial regression to evaluate the association between GLP-1RA prescriptions and AUDIT-C scores. Figure 3 displays the main effect estimates for our primary and secondary analyses, alongside a visual summary of the estimated effects and 95% CIs. In our primary analysis, after adjusting for relevant covariates, participants with active GLP-1RA prescriptions had significantly lower AUDIT-C scores compared with those who did not yet have a GLP-1RA prescription (i.e., the future prescription group). Specifically, those with active GLP-1RA prescriptions had an IRR of 0.95 (95% CI, 0.91-0.99; *P* = 0.01), indicating that having an active GLP-1RA prescription was associated with a 5% lower average AUDIT-C score.

Participants with a former GLP-1RA prescription had lower AUDIT-C scores compared with those with future prescriptions, but this effect was not statistically significant (IRR: 0.92; 95% CI, 0.85-1.01; *P* = 0.07). Models stratified by sex showed that male and female participants with active GLP-1RA prescriptions had lower AUDIT-C scores than their counterparts with future GLP-1RA prescriptions, but this association was statistically significant only among females (male IRR: 0.95; 95% CI, 0.89-1.02; *P* = 0.15; female IRR: 0.95; 95% CI, 0.91-1.00; *P* = 0.04).

We could not conduct sex-stratified analyses among the participants with former GLP-1RA prescriptions due to insufficient sample sizes.

The results of the secondary analyses, which compared participants with active and former GLP-1RA prescriptions to propensity-score matched comparison groups, were similar to the primary analysis. The active group was even lower in this analysis, at an 11% lower average AUDIT-C score (IRR: 0.89; 95% CI, 0.85-0.93; *P* = <0.001), and we found no significant difference for the former group (IRR: 0.98; 95% CI, 0.87-1.11; *P* = 0.77).

We additionally ran weighted negative binomial and weighted Poisson regressions (where appropriate) to evaluate the association between GLP-1RAs and each of the three composite AUDIT-C questions separately. Participants with former and active GLP-1RA prescriptions had significantly lower reported drinking frequency compared with future GLP- 1RA recipients (active IRR: 0.96; 95% CI, 0.92-0.99; *P* = 0.02; former IRR: 0.90; 95% CI, 0.82-0.98; *P* = 0.01). There were no significant differences for either group in the quantity of drinks per day or the number of occasions with six or more drinks. Full model estimates, including covariates, are presented in eTables 4-15.

## Discussion

Among *All of Us* participants with GLP-1RA prescriptions, we found that having an active GLP-1RA prescription was associated with lower AUDIT-C scores compared with those who had not yet received GLP-1RA prescriptions and a matched comparison group. Although the effect size was modest, the findings remain clinically and publicly relevant, given that the association was observed in a general population prescribed GLP-1RAs for non–alcohol-related indications rather than a treatment-seeking cohort. Notably, in participants who formerly had a GLP-1RA prescription, this association was not statistically significant. The latter findings are consistent with the hypothesis that, similar to weight regain after GLP-1RA discontinuation,^24^ people who reduce drinking after taking GLP-1RAs may rebound after medication is stopped; however, this remains speculative given the lack of longitudinal data.

There were minimal differences in the estimated IRRs by sex for participants with active prescriptions, although the association was significant for only female participants, likely owing to smaller sample sizes. In the analysis of the individual questions that comprise the AUDIT-C, participants with active and former GLP-1RA prescriptions reported a statistically significant but modestly lower frequency of drinking but not fewer drinks per occasion or less binge drinking.

To our knowledge, few EHR-based pharmacoepidemiological studies have examined the association between GLP-1RA prescriptions and self-reported alcohol consumption, particularly in a large, diverse, real-world sample. Although the AUDIT-C was asked at only one point, we identified a comparison group of future GLP-1RA recipients and found similar results to a matched comparison group (not required to have future GLP-1RA prescriptions). Our findings are like those of other EHR-based studies that have found associations between GLP-1RAs and reduced alcohol consumption among smaller or more constrained populations^13,14^ and studies that found reduced incidence of other alcohol-related outcomes.^8–12^ Additionally, we examined associations for participants who no longer had an active GLP-1RA prescription and found that they reported a lower frequency of alcohol consumption.

Taken together, these findings support that GLP-1RAs may reduce alcohol consumption, particularly by decreasing the frequency at which people choose to drink. The latter point is not fully consistent with a recent randomized controlled trial where semaglutide reduced the amount of alcohol intake on drinking days but did not change the overall number of drinking days.^25^ Further studies are needed to delineate what specific aspects and parameters of alcohol seeking and drinking are more amenable in response to GLP-1RAs.

By comparing with future GLP-1RA recipients and a matched comparison group, we aimed to maximize similarity between the groups to resemble randomness in RCTs. We cannot rule out the possibility that GLP-1RA recipients decreased their alcohol consumption for other reasons. GLP-1RA recipients may have been exposed to concomitant interventions or behavioral changes such as being in a weight-loss program or attempting to mitigate other health concerns (e.g., diabetes). Additionally, despite representing a wide range of people in the United States, *All of Us* cannot be generalized to the entire U.S. population given purposive recruitment efforts.^26^ The participants who took the *All of Us* Lifestyle survey may be self-selecting, and participants may choose to skip questions entirely, which could bias responses to the AUDIT-C questions. Additionally, the AUDIT-C questions included in the Lifestyle Survey do not use the common modification for women and those aged 65 or older which allows for a lower threshold for binge drinking. Thus, our results could be underestimating clinically meaningful alcohol use for these groups. Participants who took the Lifestyle survey after their GLP-1RA prescriptions (i.e., the active and former groups) were likely to be earlier GLP-1RA recipients who did not have access to the same types of GLP-1RAs or a broader array of FDA-approved uses (eTable 18).

This study, with some limitations, contributes to the growing body of literature that suggests GLP-1RAs may lower alcohol consumption, leading to fewer adverse alcohol-related health outcomes. *All of Us* represents a unique opportunity to examine these associations further by providing multimodal data, such as genomics, wearables, and other surveys alongside EHR data, to better understand these associations. Given the explosive growth of GLP-1RA uptake in the United States, the release of additional data through *All of Us*, such as the upcoming CDRv9, may offer greater power and possibilities for study. Additionally, the present findings are in line with emerging data from primary or secondary analyses of RCTs indicating that, compared with placebo, GLP-1 receptor agonism leads to reduction in alcohol drinking.^25,27–28^ Nonetheless, results from additional and larger RCTs are needed to further test the safety and efficacy of GLP- 1RAs in AUD.^29^

## Data Availability

All data in the present study is available to registered researchers through the All of Us Researcher Workbench: https://www.researchallofus.org.
The code and output is available in a workspace on the All of Us Researcher Workbench at: https://workbench.researchallofus.org/workspaces/aou-rw-103b7f16/glp1driver/analysis

## ACKNOWLEDGMENT

**Author Contributions**

Dr Tyndall, Ms Gasdaska, Mr Brannock, and Dr Preble had full access to all data in the study and take responsibility for the integrity of the data and the accuracy of the data analysis. *Concept and design:* All authors

*Acquisition,* analysis*, or interpretation of data:* All authors

*Drafting of* the *manuscript:* Tyndall, Gasdaska, Brannock, Preble, Huda

*Critical* review *of the manuscript for important intellectual content:* All authors

*Statistical* analysis: Gasdaska, Brannock

*Obtained* funding: Adjemian

*Administrative, technical, or material support:* Litwin, Sastry, Adjemian

*Supervision:* McPheeters, Marcial, Adjemian

## Additional Contributions

We gratefully acknowledge *All of Us* participants for their contributions, without whom this research would not have been possible. We also thank the National Institutes of Health’s *All of Us* Research Program for making available the participant data examined in this study.

We thank Karen Kesler, PhD, for advising on methodology, Jessica Cance, MPH, PhD, for reviewing a draft, Sheryl Cates for project administration and Jennifer D. Uhrig, PhD, and Megan A. Lewis, PhD, the award multiple principal investigators, for supporting this project (RTI International).

## Data and code Availability

The Featured Workspace is hosted on the *All of Us* Researcher Workbench and contains code and data used for the analysis in the paper: https://workbench.researchallofus.org/workspaces/aou-rw-103b7f16/glp1driver/analysis

