## Supplemental Materials for "Association of GLP-1 Receptor Agonist Prescriptions and Alcohol Consumption in the National Institutes of Health’s *All of Us* Cohort"

SUPPLEMENT

Contents

Note on tobacco and other substance use	1

eFigure 1. CONSORT Diagram Showing Attrition from Exclusion Criteria for Matched Comparison Groups	1

eFigure 2. Covariate Balance in Active GLP-1RA Group Before and After Inverse Probability of Treatment Weighting with Future GLP-1RA Group	2

eFigure 3. Covariate Balance in Former GLP-1RA Group Before and After Inverse Probability of Treatment Weighting with Future GLP-1RA Group	3

eFigure 4. Covariate Balance in Active GLP-1RA Group Before and After Propensity Score Matching with Matched Comparison Group	4

eFigure 5. Covariate Balance in Former GLP-1RA Group Before and After Propensity Score Matching with Matched Comparison Group	5

eTable 1. OMOP Concept IDs Used to Define Any EHR-Derived Features	6

eTable 2. OMOP Concept IDs Used to Define GLP-1RA Exposures	8

eTable 3. ICD-9 and ICD-10 Codes for Calculating Modified Charlson Comorbidity Index	8

eTable 4. Charlson Comorbidity Index Scoring System	8

eTable 5. Charlson Comorbidity Index Hierarchy Categories2	8

eTable 6. Negative Binomial Model Evaluating Association of Alcohol Use (AUDIT-C) with Active GLP-1RA Prescriptions Compared to Future GLP-1RA Prescriptions: Primary Analysis	8

eTable 7. Negative Binomial Model Evaluating Association of Alcohol Use (AUDIT-C) with Active GLP-1RA Prescriptions Compared to Future GLP-1RA Prescriptions: Female Sex Primary Analysis	8

eTable 8. Negative Binomial Model Evaluating Association of Alcohol Use (AUDIT-C) with Active GLP-1RA Prescriptions Compared to Future GLP-1RA Prescriptions: Male Sex Primary Analysis	8

eTable 9. Negative Binomial Model Evaluating Association of Alcohol Use (AUDIT-C) with Former GLP-1RA Prescriptions Compared to Future GLP-1RA Prescriptions: Primary Analysis	8

eTable 10. Negative Binomial Model Evaluating Association of Alcohol Use (AUDIT-C) with Active GLP-1RA Prescriptions Compared to Matched Group: Secondary Analysis	8

eTable 11. Negative Binomial Model Evaluating Association of Alcohol Use (AUDIT-C) with Former GLP-1RA Prescriptions Compared to Matched Group: Secondary Analysis	8

eTable 12. Negative Binomial Model Evaluating Association of Alcohol Use with Active GLP-1RA Prescriptions Compared to Future GLP-1RA Prescriptions: AUDIT-C Drink Frequency Question Analysis	8

eTable 11. Negative Binomial Model Evaluating Association of Alcohol Use with Active GLP-1RA Prescriptions Compared to Future GLP-1RA Prescriptions: AUDIT-C Drink Quantity Question Analysis	8

eTable 13. Negative Binomial Model Evaluating Association of Alcohol Use with Active GLP-1RA Prescriptions Compared to Future GLP-1RA Prescriptions: AUDIT-C Binge Drinking Question Analysis	8

eTable 14. Negative Binomial Model Evaluating Association of Alcohol Use with Former GLP-1RA Prescriptions Compared to Future GLP-1RA Prescriptions: AUDIT-C Drink Frequency Question Analysis	8

eTable 14. Negative Binomial Model Evaluating Association of Alcohol Use with Former GLP-1RA Prescriptions Compared to Future GLP-1RA Prescriptions: AUDIT-C Drink Quantity Question Analysis	8

eTable 15. Negative Binomial Model Evaluating Association of Alcohol Use with Former GLP-1RA Prescriptions Compared to Future GLP-1RA Prescriptions: AUDIT-C Binge Drinking Question Analysis	8

eTable 18. GLP-1RA Types for the Active, Former and Future Prescription Groups	8

REFERENCES	8

### **Note on tobacco and other substance use**

Participants were categorized as having a history of tobacco use if they indicated any current or former use of cigarettes, electronic nicotine products, cigars, hookah, or smokeless tobacco products. Participants were categorized as having a history of substance use if they indicated ever using marijuana; cocaine; prescription stimulants for nonmedical reasons; other stimulants like methamphetamine, inhalants, sedatives, or sleeping pills for nonmedical reasons; hallucinogens; illicitly obtained opioids; or prescription opioids used for nonmedical reasons.^1^

### **eFigure 1. CONSORT Diagram Showing Attrition from Exclusion Criteria for Matched Comparison Groups**

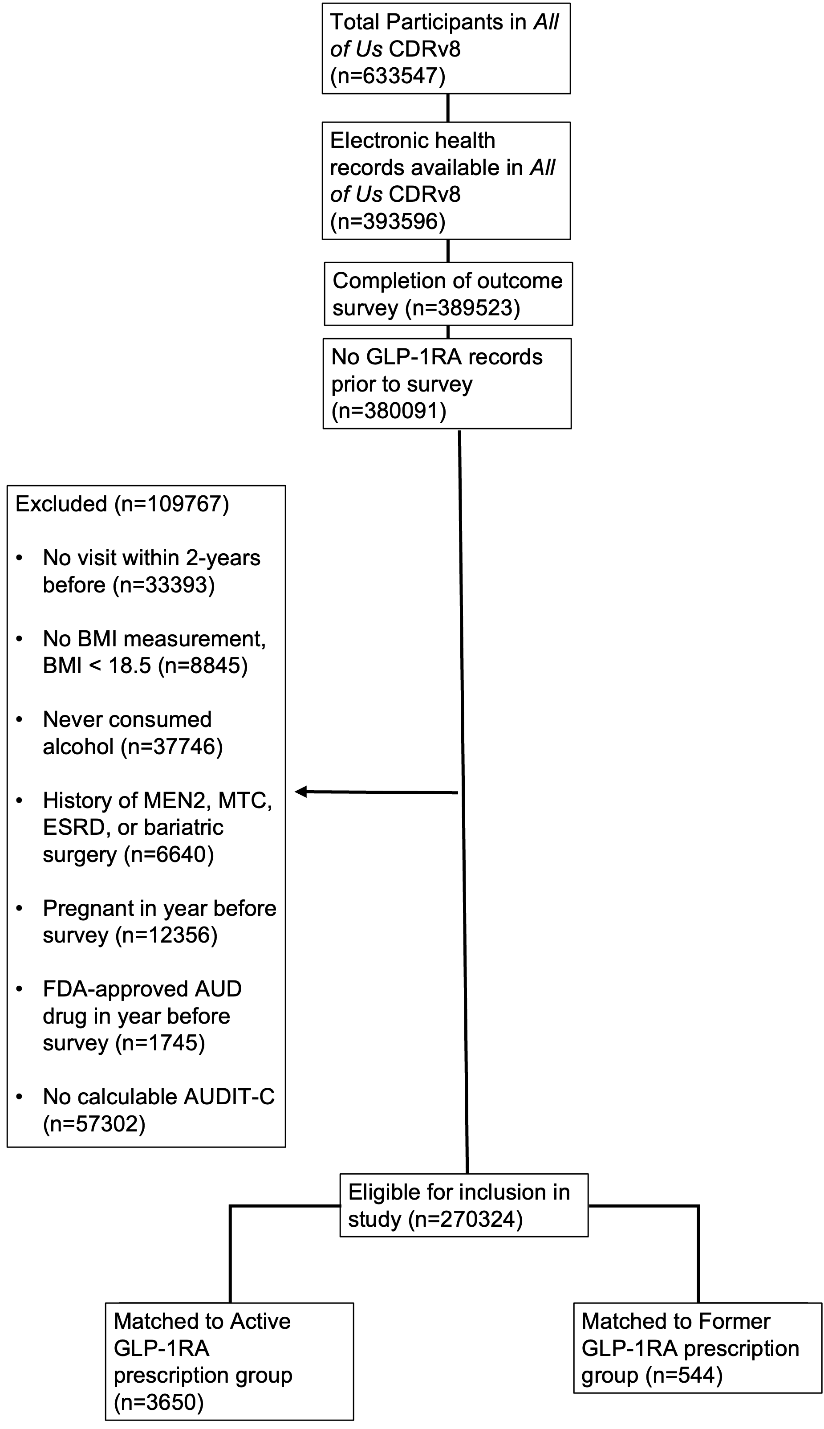

### **eFigure 2. Covariate Balance in Active GLP-1RA Group Before and After Inverse Probability of Treatment Weighting with Future GLP-1RA Group**

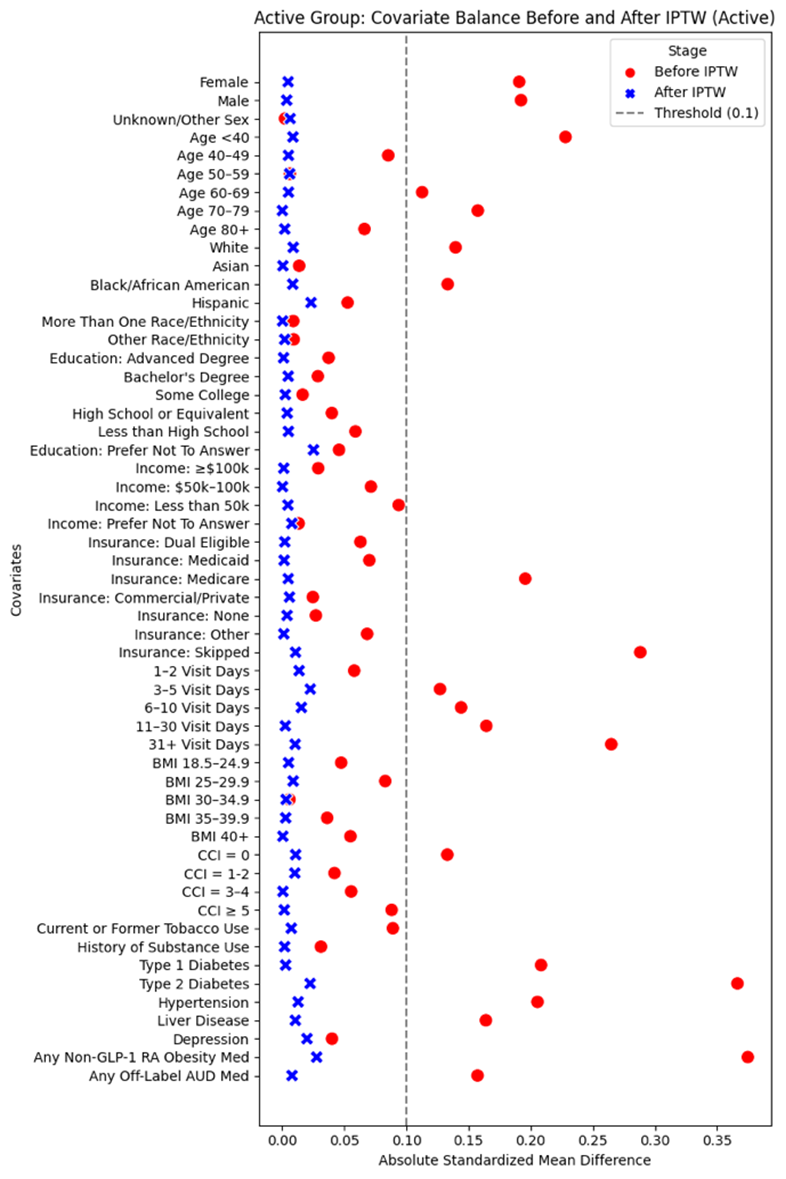

Abbreviations: AUD, alcohol use disorder; BMI, body mass index; CCI, Charlson Comorbidity Index; GLP-1RA, glucagon-like peptide-1 receptor agonist; IPTW, inverse probability of treatment weighting.

### **eFigure 3. Covariate Balance in Former GLP-1RA Group Before and After Inverse Probability of Treatment Weighting with Future GLP-1RA Group**

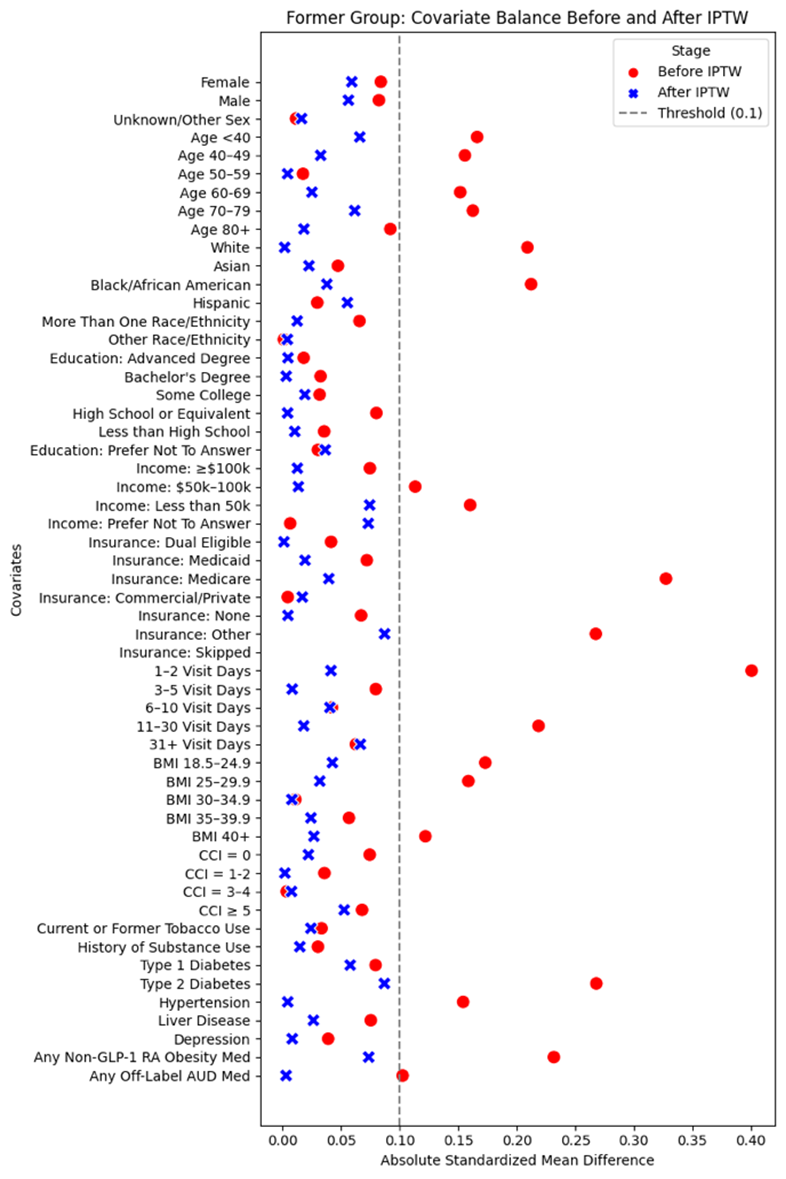

Abbreviations: AUD, alcohol use disorder; BMI, body mass index; CCI, Charlson Comorbidity Index; GLP-1RA, glucagon-like peptide-1 receptor agonist; IPTW, inverse probability of treatment weighting.

### **eFigure 4. Covariate Balance in Active GLP-1RA Group Before and After Propensity Score Matching with Matched Comparison Group**

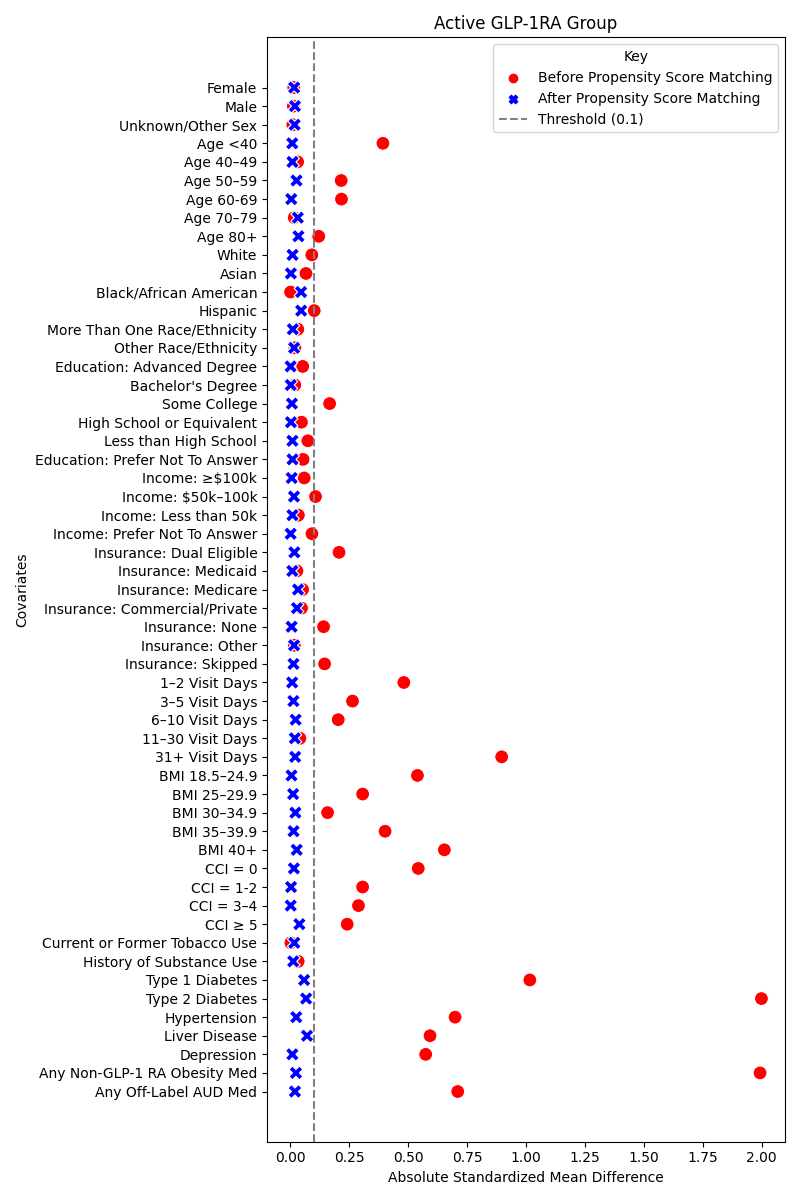

Abbreviations: AUD, alcohol use disorder; BMI, body mass index; CCI, Charlson Comorbidity Index; GLP-1RA, glucagon-like peptide-1 receptor agonist; IPTW, inverse probability of treatment weighting.

### **eFigure 5. Covariate Balance in Former GLP-1RA Group Before and After Propensity Score Matching with Matched Comparison Group**

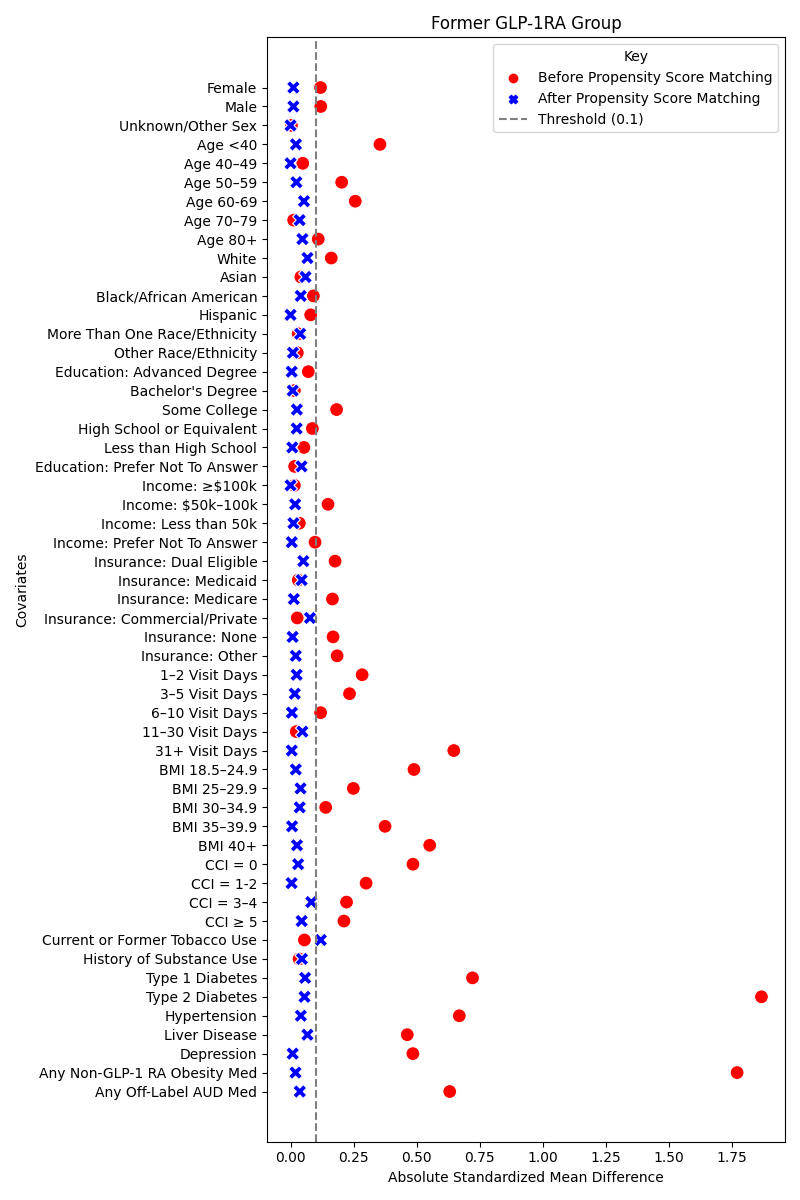

Abbreviations: AUD, alcohol use disorder; BMI, body mass index; CCI, Charlson Comorbidity Index; GLP-1RA, glucagon-like peptide-1 receptor agonist; IPTW, inverse probability of treatment weighting.

### **eTable 1. OMOP Concept IDs Used to Define Any EHR-Derived Features**

| Visit type |  |
| --- | --- |
| Inpatient visits | 42898160, 8920, 38004311, 38004284, 38004303, 8971, 8546, 32037, 8676, 38004515, 38004285, 262, 581379, 8717, 4313303, 9201 |
| Outpatient visits | 38004250, 38004251, 38004207, 8756, 38004258, 581475, 38004249, 38004696, 8964, 8966, 38004245, 581476, 38004218, 38004228, 38004238, 581479, 38004259, 43527986, 38004264, 8949, 38004222, 43527904, 38004269, 38004208, 38004225, 8883, 581385, 38004247, 38004262, 5083, 38004268, 38004267, 38004519, 38004677, 8782, 581477, 8947, 38004246, 38004227, 9202 |
| Emergency department visits | 4163685, 9203, 8870 |
| Condition |  |
| Type 1 diabetes | 443412, 201254, 37016348, 40484648, 200687, 377821, 45769876, 435216, 37017431, 42538169, 4227210, 37016767, 318712, 45763584, 37016179, 439770, 45770902, 43531008, 45769832, 4225656, 37016180, 4099214, 45769830, 4223303, 4145827, 4225055, 45773688, 45763583, 4224254, 4228112, 4152858, 4063042, 37312218, 201531, 45769873, 609120, 45757674, 35626069, 443592, 609098, 37018566, 609118, 4143857, 609115, 45757507, 45757362, 609100, 45757432, 36713094, 603320, 4047906, 43531009, 603319, 609102, 37017429, 603318, 45771075, 37016353, 37312201, 45773576, 43530660, 45757535, 43531006, 609070, 45771067, 37312200, 4102018, 609111, 43531565, 609110, 45757074, 45769901, 45769903, 4099215 |
| Type 2 diabetes | Condition concept IDs:  4193704, 201826, 37016349, 376065, 43531578, 37017432, 443729, 443731, 443732, 45757363, 443733, 45757435, 43530690, 4221495, 37016768, 40482801, 43531616, 43530685, 4099651, 45770830, 45770881, 4222876, 43531563, 4222415, 443734, 4130162, 43530656, 37016354, 45757474, 4196141, 201530, 4226121, 4304377, 4200875, 43531564, 36714116, 4140466, 45773064, 45770880, 4063043, 45771064, 35626070, 609103, 43530689, 43531653, 4228443, 45757449, 4230254, 43531651, 36712687, 609117, 609099, 609106, 36712686, 609101, 4099216, 45769905, 37312204, 609119, 609116, 4215719, 43531577, 37018728, 4177050, 609104, 4129519, 609109, 609105, 602345, 37312205, 43531597, 46274058, 45757278, 45769888, 45757499, 609112, 45757277, 45757280, 4321756, 43531566, 43531562, 43531588, 43531559, 45769906, 4221487, 37018912, 4130164, 45769875, 45770831, 45757445, 45757446, 43531608  Drug concept IDs:  40164929, 40164897, 40163924, 1503297, 1525221, 19077638, 40164930, 19077636, 45774754, 45774442, 793152, 45774758, 40164898, 40163926, 45774893, 45774448, 19078552, 40169222, 19125043, 40169223, 40164946, 793147, 45774446, 45774438, 45774895, 1560171, 19125041, 1597772, 793143, 19079990, 19077682, 1597773, 37003617, 19080793, 1597761, 40163928, 19079293, 45892686, 19125049, 37003616, 19125051, 40239219, 19077637, 44785831, 37003607, 40170911, 19078551, 741834, 40220373, 45774435, 793154, 19078603, 19077659, 19006932, 1537597, 45774751, 1580747, 44785473, 780249, 40220376, 1559684, 40239218, 40166816, 19078530, 19030445, 43013924, 780260, 1525220, 1596916, 43526467, 46234237, 780256, 1597757, 44785477, 19009384, 46221581, 44785833, 1525216, 40167020, 40166814, 43526468, 1525215, 40220375, 19030443, 40167021, 1537599, 1525217, 19125045, 19125047, 1597756, 1583722, 1537596, 1597760, 37003609, 793153, 44785829, 37496842, 40164925, 40164947, 1596960, 1537601, 1559685, 40220371, 40164892, 40163929, 40166041, 40166815, 780266, 1537603, 37496746, 1597759, 741832, 37496844, 37496840, 43526472, 40164891, 19023424, 43013911, 45774533, 1596957, 779705, 19023425, 1537605, 1583729, 43526471, 1537598, 37003606, 19012270, 19023063, 45774531, 40163927, 1586345, 19079465, 780262, 1503328, 1537602, 1525218, 40166002, 40166037, 1547504, 780248, 1583730, 40163922, 1537600, 780259, 19125223, 19023426, 1547510, 40165997, 19006910, 1529352, 1583727, 1537604, 40163923, 37003608, 19027257, 1518148, 40239216, 40163925, 1503327, 780255, 37496838, 1529331, 793114, 42708171, 40164923, 19135405, 43526465, 19054444, 780264, 19054443, 42708078, 42708079, 40164932, 1596956, 19125221, 40166042, 1516766, 793111, 40164882, 37499746, 43013928, 793327, 42708167, 1516771, 19054445, 1547506, 1502830, 780265, 19135263, 40167017, 37496832, 1560168, 780258, 19047663, 19021312, 1516770, 42708170, 40164922, 40164900, 40166003, 19047612, 780261, 40164881, 44816273, 793331, 40165975, 40167016, 46287691, 42708087, 40165972, 43013884, 1718604, 19125219, 43013925, 19006933, 40165998, 793329, 42708166, 46287410, 1718711, 793321, 19126618, 1592795, 1560233, 1502829, 19099245, 1547505, 1502826, 1547508, 42903113, 40164943, 19081295, 40164915, 19047638, 1718599, 1559784, 1592794, 19040969, 42708175, 44816274, 19117945, 40166038, 1502827, 45892179, 19099207, 19023585, 19099208, 19071495, 40164904, 40164894, 40166035, 40164944, 44816282, 45775999, 780263, 1559782, 1594973, 46287408, 45892176, 40063354, 40165994, 793293, 42708086, 42708174, 19099209, 1547507, 45775621, 45774714, 19030580, 1592800, 19117200, 45775455, 42708089, 1592798, 19001408, 19099246, 43013912, 1592804, 1592803, 44816283, 42902992, 40165976, 43013896, 780257, 40164948, 45774721, 40164905, 45775860, 1718706, 40165973, 45892172, 19030579, 40167636, 1592653, 46287689, 1592656, 46234234, 19077684, 45774722, 19025946, 40164938, 19040970, 19077681, 46287684, 1515107, 45774710, 44816332, 40168374, 40229046, 19120648, 19030575, 19062999, 40229045, 1515249, 36249798, 19085004, 40164942, 45892177, 45774719, 40165995, 1596918, 42708088, 40164917, 42902823, 19117198, 1559760, 40167022, 40164889, 40164886, 42903382, 37497424, 19120315, 36249793, 19134639, 40229047, 19010532, 43013929, 19010204, 46287688, 43013899, 42708091, 43013897, 40171448, 41348912, 40165970, 40229048, 40165969, 19030572, 46287686, 40044251, 45774724, 37496783, 46287680, 40164913, 19078029, 19078030, 40063350, 45775867, 21110724, 19027259, 19121940, 793300, 42903059, 40164916, 45775623, 19012702, 40229050, 19078028, 19030576, 40164907, 19125042, 1597775, 793306, 19086484, 19045606, 40164888, 43013915, 37499835, 19012703, 21051793, 43013918, 793313, 19006908, 40164920, 19078607, 1718899, 1718901, 1502811, 43013900, 40164880, 45774717, 37496785, 40168370, 40168375, 1597792, 19060409, 19047829, 1510206, 40164941, 19029029, 19022851, 40164910, 40171449, 19047828, 1502809, 40166004, 21159907, 40166044, 43013916, 44057249, 19088818, 42708090, 1547554, 37499837, 1594976, 19029030, 1597776, 40164914, 45776000, 19045630, 19021786, 1525242, 1510207, 40164919, 19120257, 1718905, 40164885, 19079986, 40171451, 793828, 1718908, 40164918, 40220862, 19122368, 40164903, 36249791, 43013919, 45774715, 40167637, 40139022, 1515250, 19082950, 40099898, 793309, 19078604, 37499839, 1718904, 40231396, 40167012, 1597795, 19078553, 19045629, 1515254, 19125040, 1597774, 19024509, 1718912, 40168371, 1525244, 40164911, 793826, 19021787, 45775619, 40229049, 19117199, 19045625, 43013921, 40164924, 40046201, 19045626, 40167013, 19134624, 1515251, 19134642, 19135386, 19081296, 1510208, 19120166, 1718898, 19024508, 19099070, 1597798, 1529358, 19006909, 40168368, 43013905, 1525243, 19030573, 1597799, 40167634, 1597801, 19116559, 1596974, 40221118, 1592323, 19029061, 40239222, 42902866, 1502812, 43013902, 43011498, 21140107, 1596973, 19024503, 45775865, 45775622, 1502810, 40168369, 43013903, 40164890, 21149954, 42901620, 1529359, 1529360, 36249796, 42901622, 1596959, 43011499, 40170912, 1594975, 21169699, 1515258, 19082961, 19060491, 1718909, 1559785, 40059665, 44506754, 19122367, 46234240, 40165977, 19024507, 19024510, 1516794, 40231393, 1515253, 19099055, 1597781, 1559761, 1559728, 40164884 |
| Depression | 440383, 4282096, 4077577, 433440, 436665, 4282316, 444100, 442306, 36684319, 44784632, 435220, 432883, 4228802, 4049623, 4195572, 439254, 438998, 441534, 432876, 4307956, 4338031, 434911, 4098302, 4244078, 4205002, 437528, 444038, 440078, 438406, 4239471, 439253, 432866, 4224940, 4152280, 443906, 435226, 439248, 443237, 4242733, 4328217, 4150985, 439256, 433992, 432285, 35622934, 439249, 442570, 441834, 4012869, 4314692, 4149320, 439246, 4105930, 372599, 440696, 443797, 436386, 439251, 4151170, 438727, 4092239, 40481798, 43531624, 439255, 37018688, 37396201, 4298317, 4329560, 4149321, 441836, 36717092, 4103574, 36713698, 4154309, 437249, 607543, 4307111, 435520, 35610112, 35624743, 4103853, 42872411, 4280361, 4037669, 4102936, 4333678, 440980, 4307804, 4220617, 37016718, 35624748, 42872722, 437250, 439259, 4287544, 4336957, 440067, 432290, 4030856, 35624747, 42872413, 433751, 4215917, 438119, 4141292, 439785, 4094358, 4250023, 439250, 443864, 4172156, 35624744, 439272, 35615153, 4029464, 440698, 4028027, 4269143, 438405, 439273, 607540, 4324945, 37111697, 440079, 35624745, 4327337, 4262111, 44782943, 35615152, 4114950, 37109941, 439262, 4102603, 439001, 4155798, 4241158, 42538589, 373176, 36714998, 4175329, 4205471, 37018689, 4185096, 4057218, 4174987, 4223090, 37312578, 4144519, 43020451, 37016268, 36714389, 4001733, 4071442, 4161200, 4154283, 4304140, 762060, 36715000, 4237734, 4129184, 4031328, 4144233, 37110429, 42538590, 4307951, 4243308, 4096229, 4133073, 4226155, 4307518, 4141603, 4197669, 4154391, 437532, 4154805, 4103126, 4148934, 4217940, 4224639 |
| Liver disease | 4059290, 194984, 201613, 4064161, 194990, 40484532, 4245975, 194692, 196463, 4267417, 200763, 46269816, 4340383, 193256, 201612, 4029488, 4337543, 201343, 4340390, 46269835, 4340386, 4055224, 4026131, 4059297, 377604, 4026125, 4026136, 199867, 4058694, 4058695, 4342883, 4340385, 4212540, 4340941, 4340948, 201065, 4243475, 4340394, 40482277, 4026032, 45769564, 46269836, 46273476, 46269818, 37396401, 4059298, 4055223, 4058696, 46269814, 4059299, 4318541, 4055210, 4238978, 36716710, 4030392, 4052963, 4159158, 4055207, 37017265, 4157032 |
| Hypertension | 320128, 312648, 44782429, 444101, 316866, 439696, 319034, 44784439, 443919, 319826, 44784621, 376965, 317898, 4110948, 44782690, 313502, 314378, 4028741, 44784638, 312938, 442604, 44782728, 439698, 314369, 195556, 43021852, 43020455, 442626, 314958, 45771064, 439695, 442603, 439694, 4289933, 316994, 37018886, 4263504, 318437, 37208172, 4209293, 4322893, 4167358, 44782689, 37311148, 44782691, 4159755, 4049389, 43021835, 45757140, 4276511, 46273164, 4058987, 44784639, 760850, 44782692, 4183981, 4263067, 43020457, 4135463, 4178312, 4322735, 45757445, 45757446, 4180283, 45771067, 42538697, 4262182, 46273636 |
| Medullary thyroid cancer | 4111011 |
| Multiple endocrine neoplasia type 2 | 24612, 36717452, 199775 |
| End-stage renal disease | 193782, 443611, 44782690, 44784638, 4030520, 43020455, 37018886, 37017813, 45768813, 4125970, 43531562, 760850, 46273164, 45769906, 45769903, 37018761 |
| Bariatric surgery | 40756884, 2108974, 2749004, 4143852, 2108990, 2748983, 40483096, 2108986, 2109002, 46270974, 2108967, 2108975, 2108966, 2108993, 4148762, 4069249, 2108988, 2109003, 2109004, 2108965, 2108987, 2749219, 2109001, 4190525, 2108989, 2749005, 2109013, 2748981, 2109000, 2748989, 2749003, 2748982, 2749002, 4171187, 2749204, 4087760, 2748429, 2108999, 2109015, 2748999, 2748984, 2109384, 4145504, 2749207, 2748994, 2749209, 2748724 |
| Obesity treatments and non-FDA-approved AUD treatment OMOP concept IDs | |
| Baclofen | 715235, 715236, 715233, 19026067, 19107271, 19010804, 715239, 19026066, 19082434, 715276, 19108242, 702368, 1759177, 40009903, 702377, 702374, 19117731 |
| Gabapentin | 19077548, 19077547, 19077549, 797399, 19077550, 19006232, 19006231, 19077571, 19006233, 797414, 797415, 40241280, 19068334, 797439, 797437, 19038876, 40221154, 797438, 40241279, 19120656, 19038875, 40221155, 40239097, 797432, 40239096, 19122310, 44785042, 797440, 19038877, 44785041, 19122313, 40241453, 19120657, 797434, 797421, 19122315, 40241452, 797433, 37003569, 1302153, 797435, 1302155, 40058892, 40845890, 37003568 |
| Varenicline | 19130125, 780446, 780442, 19127968, 19124311, 19130124, 780444, 19128025, 780447, 19089147, 702092, 702086, 19089146, 780445 |
| Topiramate | 742303, 19022457, 742268, 742304, 742267, 40172928, 742302, 40172930, 742269, 40172924, 742270, 19028938, 42873601, 42873596, 40172926, 42873600, 42873597, 19028939, 742271, 742344, 742345, 19116968, 42901669, 43560435, 43560429, 43560433, 43560430, 43560436, 42901670, 19082967, 42873604, 19082968, 43560432, 1718081, 1718080, 42873605, 43560431, 1718082, 43560434, 44816192, 1718079, 44816200, 19082969, 40901806, 44816190, 44816196, 742343, 742311, 44816198, 19082981, 44816193, 42901672, 43560759, 19082009, 43189984, 742309, 702345, 19082549, 42901671 |
| Prazosin | 1350521, 1350552, 19040227, 1350490, 19040228, 1350489, 19040229, 40239424, 19012595, 19085488, 1350553, 19012596, 1350554, 40078731, 19111604, 948855 |
| Ondansetron | 19019419, 19005965, 1000560, 19079171, 19005966, 40168116, 19079172, 19005967, 40168118, 19000634, 19083034, 40223993, 19000635, 40168119, 40223992, 1000612, 40070107, 43203720, 1000616, 19079159, 19068361, 40223995, 19117094, 40223996, 19085099, 40070098, 19000639 |
| Phentermine | 19131610, 40176158, 735340, 19131607, 40173346, 19131611, 735510, 735533, 42873601, 42873596, 735511, 42873600, 42873597, 40220687, 42901669, 42901670, 19131609, 40239828, 42873604, 42873605, 42708885, 735431, 735473, 735534, 40239831, 42873384, 735422, 735476, 735479, 735477, 19133190, 19131606, 19134205, 40239829, 19131608, 735475, 19043400, 19133202, 735507, 40239832, 19133182, 19133184, 42901672, 735423, 735480, 19043421, 735502, 735481, 735504, 42901671, 735474, 42708886, 735506, 735505, 735394 |
| Phendimetrazine | 40176940, 40182492, 40222539, 723344, 37499822, 40221844, 40176941, 40182493 |
| Diethylpropion | 40177103, 40176755, 725822, 40176763, 40177105, 40176774, 40177106, 40177104 |
| Orlistat | 741572, 741530, 741575, 741531, 741576, 19120531, 19085100, 19012271 |
| Metformin | 40164929, 40164897, 40163924, 1503297, 40164930, 40164898, 40163926, 40164946, 40163928, 40220373, 19006932, 40220376, 40220375, 40164925, 40164947, 40220371, 40164892, 40163929, 40164891, 40163927, 1503328, 40166002, 40163922, 40165997, 40163923, 19027257, 40163925, 1503327, 42708171, 40164923, 40164932, 40164882, 42708167, 42708170, 40164900, 40164922, 40166003, 40164881, 40165975, 42708087, 46287691, 40165972, 40165998, 19006933, 42708166, 46287410, 1592795, 40164943, 40164915, 1592794, 42708175, 40164904, 40164894, 45775999, 40164944, 40063354, 46287408, 42708086, 42708174, 40165994, 45775621, 45774714, 1592800, 42708089, 45775455, 1592798, 40165976, 43013896, 1592804, 1592803, 45775860, 45774721, 40164905, 40164948, 40165973, 45774722, 46287689, 1592656, 1592653, 46287684, 40164938, 40229046, 45774710, 40164942, 36249798, 40229045, 42708088, 40165995, 40164917, 45774719, 40164889, 36249793, 40164886, 40229047, 42708091, 43013897, 43013899, 46287688, 46287686, 40165969, 40229048, 40165970, 40171448, 21110724, 40063350, 45775867, 45774724, 46287680, 40164913, 37496783, 19086484, 793306, 40164907, 19121940, 19027259, 40164916, 40229050, 793300, 45775623, 1718901, 40164880, 43013900, 793313, 40164920, 1718899, 21051793, 40164888, 37499835, 40164910, 40171449, 40164941, 37496785, 45774717, 40171451, 793828, 1718908, 40164918, 40164885, 40164919, 1718905, 40164914, 45776000, 40166004, 42708090, 37499837, 19088818, 21159907, 40164924, 45775619, 40229049, 1718904, 40231396, 1718912, 40164911, 793826, 40164903, 45774715, 37499839, 36249791, 793309, 1718909, 21169699, 40164884, 36249796, 40231393, 40165977, 45775622, 21140107, 45775865, 21149954, 40164890, 1718898 |
| FDA-approved AUD drugs | 44784878, 1714319, 19043981, 1714372, 45774490, 44784880, 45774486, 44784879, 1714351, 735851, 19043982, 19043959, 19044538, 21106133, 735850, 735882, 40063517, 40166395, 40175986, 40166398, 19044893, 40166399, 19082217, 40166396, 19020355, 735887, 40166401, 40166403, 40166404, 735888, 40166392, 19119755, 40166400, 40166393, 40166407, 40166408 |

Abbreviations: AUD, alcohol use disorder; EHR, electronic health record; FDA, U.S. Food and Drug Administration; OMOP, Observational Medical Outcomes Partnership.

### **eTable 2. OMOP Concept IDs Used to Define GLP-1RA Exposures**

| **GLP-1RA** | **OMOP concept ID** | **RxNorm code** |
| --- | --- | --- |
| Albiglutide | 44816332 | 1534763 |
| Dulaglutide | 45774435 | 1551291 |
| Exenatide | 1583722 | 60548 |
| Liraglutide | 40170911 | 475968 |
| Lixisenatide | 44506754 | 1440051 |
| Semaglutide | 793143 | 1991302 |
| Tirzepatide | 779705 | 2601723 |

Abbreviations: GLP-1RA, glucagon-like peptide-1 receptor agonist; OMOP, Observational Medical Outcomes Partnership.

### **eTable 3. ICD-9 and ICD-10 Codes for Calculating Modified Charlson Comorbidity Index**

| **Condition** | **ICD-9 diagnosis codes** | **ICD-10 diagnosis codes** |
| --- | --- | --- |
| Myocardial infarction | 410, 412, 410.x, 412.x, | I21, I22, I21.x, 122.x, 125.2 |
| Congestive heart failure | 398.91, 402.01, 402.11, 402.91, 404.01, 404.03, 404.11, 404.13, 404.91, 404.93, 425.4, 425.5, 425.6, 425.7, 425.8, 425.9, 428 | I11.0, I13.0, I13.2, I25.5, I42.0, I42.5, I42.6, I42.7, I42.8, I42.9, P29.0, I43, 150, I43.x, I50.x |
| Peripheral vascular disease | 093.0, 437.3, 443.1, 443.9, 447.1, 557.1, 557.9, V43.4, 440, 441, 440.x, 441.x, 443.2x, 443.8x | I73.1, I73.8, I73.9, I77.1, I79.0, I79.1, I79.8, K55.1, K55.8, K55.9, Z95.8, Z95.9, I70, I71, I70.x, I71.x |
| Cerebrovascular disease | 362.34, 430, 431, 432, 433, 434, 435, 436, 437, 438, 430.x, 431.x, 432.x, 433.x, 434.x, 435.x, 436.x, 437.x, 438.x | G45, G46, I60, I61, I62, I63, I64, I65, I66, I67, I68, G45.x, G46.x, G32.0x, H34.0x, H34.1x, H34.2x, I60.x, I61.x, I62.x, I63.x, I64.x, I65.x, I66.x, I67.x, I68.x |
| Dementia | 290.0, 290.3, 294.0, 294.8, 331.0, 331.2, 331.7, 797, 290.1x, 290.2x, 290.4x, 294.1x, 294.2x, 331.1x | F04, F05, F06.1, F06.8, G13.2, G13.8, G31.1, G31.2, G91.4, G94, R41.81, R54, F01, F02, F03, G30, F01.x, F02.x, F03.x, G30.x, G31.0x |
| Chronic pulmonary disease | 506.4, 508.1, 508.8, 490, 491, 492, 493, 494, 495, 496, 500, 501, 502, 503, 504, 505, 490.x, 491.x, 492.x, 493.x, 494.x, 495.x, 496.x, 500.x, 501.x, 502.x, 503.x, 504.x, 505.x | J68.4, J70.1, J70.3, J40, J41, J42, J43, J44, J45, J46, J47, J60, J61, J62, J63, J64, J65, J66, J67, J40.x, J41.x, J42.x, J43.x, J44.x, J45.x, J46.x, J47.x, J60.x, J61.x, J62.x, J63.x, J64.x, J65.x, J66.x, J67.x |
| Rheumatic disease | 446.5, 710.0, 710.1, 710.2, 710.3, 710.4, 714.0, 714.1, 714.2, 725, 714.8x, 725.x | M31.5, M35.1, M35.3, M36.0, M05, M06, M32, M33, M34, M05.x, M06.x, M32.x, M33.x, M34.x |
| Peptic ulcer | 531, 532, 533, 534, 531.x, 532.x, 533.x, 534.x | K25, K26, K27, K28, K25.x, K26.x, K27.x, K28.x |
| Renal disease, mild to moderate | 403.00, 403.10, 403.90, 404.00, 404.01, 404.10, 404.11, 404.90, 404.91, 585.1, 585.2, 585.3, 585.4, 585.9, V42.0, 582, 583, 582.x, 583.x | I12.9, I13.0, I13.10, N18.1, N18.2, N18.3, N18.4, N18.9, Z94.0, N03, N05, N03.x, N05.x |
| Hemiplegia or paraplegia | 334.1, 342, 343, 344, 342.x, 343.x, 344.x | G04.1, G11.4, G80.0, G80.1, G80.2, G81, G82, G83, G81.x, G82.x, G83.x |
| Any malignancy | 180 – 195, 200 – 208, 199.1, 238.6, 180.x – 195.x, 200.x – 208.x, | C00 - C34, C37 – C41, C43, C45 – C58, C60 - C63, C76, C80.1, C81 - C85, C88, C90 - C99, C00.x - C34.x, C37.x - C41.x, C43.x, C45.x - C58.x, C60.x - C63.x, C76.x, C81.x - C85.x, C88.x, C90.x - C99.x, |
| Renal disease, severe | 403.01, 403.11, 403.91, 404.02, 404.03, 404.12, 404.13, 404.92, 404.93, 585.5, 585.6, 588.0, V45.11, V45.12, V56.0, V56.1, V56.2, V56.31, V56.32, V56.8, 586, 586.x | I12.0, I13.11, I13.2, N18.5, N18.6, N25.0, Z99.2, N19, Z49, N19.x, Z49.x |
| HIV infection, no AIDS | 042 - 044 | B20-B22, B24 |
| Metastatic solid tumor | 196 – 198, 199.0, 196.x - 198.x | C77-C79, C80.0, C80.2, C77.x - C79.x |
| AIDS (HIV infection + opportunistic infection) | 117.5, 007.4, 078.5, 007.2, 136.3, V12.61, 046.3, 003.1, 799.4, 112, 180, 114, 348.3, 054, 115, 176, 031, 130, 200, 201, 202, 203, 204, 205, 206, 207, 208, 010 - 018, 112.x, 180.x, 114.x, 348.3.x, 054.x, 115.x, 176.x, 031.x, 130.x, 200.x, 201.x, 202.x, 203.x, 204.x, 205.x, 206.x, 207.x, 208.x, 010.x - 018.x, | B37, C53, B38, B45, B25, G93.4, B39, C46, A31, B58, C81 - C96, A15, A16, A17, A18, A19, A07.2, B00, A07.3, B59, Z87.01, A81.2, A02.1, R64, B37.x, C53.x, B38.x, B45.x, B25.x, G93.4x, B39.x, C46.x, A31.x, B58.x, C81.x - C96.x, A15.x, A16.x, A17.x, A18.x, A19.x |

Abbreviations: ICD, International Classification of Diseases.

### **eTable 4. Charlson Comorbidity Index Scoring System**

| **Condition number** | **Condition description** | **Points** |
| --- | --- | --- |
| 1 | Myocardial infarction | 1 |
| 2 | Congestive heart failure | 1 |
| 3 | Peripheral vascular disease | 1 |
| 4 | Cerebrovascular disease | 1 |
| 5 | Dementia | 1 |
| 6 | Chronic pulmonary disease | 1 |
| 7 | Rheumatic disease | 1 |
| 8 | Peptic ulcer | 1 |
| 9 | Renal disease, mild to moderate | 1 |
| 10 | Hemiplegia or paraplegia | 2 |
| 11 | Any malignancy | 2 |
| 12 | Renal disease, severe | 3 |
| 13 | HIV infection, no AIDS | 3 |
| 14 | Metastatic solid tumor | 6 |
| 15 | AIDS | 6 |

### **eTable 5. Charlson Comorbidity Index Hierarchy Categories^2^**

| **Category** | **Hierarchy rule** |
| --- | --- |
| 1 | Hemiplegia/paraplegia (Condition 10) trumps cerebrovascular disease (Condition 4) |
| 2 | Renal disease, severe (Condition 12) trumps renal disease, mild to moderate (Condition 9) |
| 3 | Metastatic solid tumor (Condition 14) trumps any malignancy (Condition 11) |
| 4 | AIDS (Condition 15) trumps HIV (Condition 13) |

Note. Within each hierarchy, the milder condition does not contribute to the Charlson Comorbidity Index score if the more severe condition is present.

### **eTable 6. Negative Binomial Model Evaluating Association of Alcohol Use (AUDIT-C) with Active GLP-1RA Prescriptions Compared to Future GLP-1RA Prescriptions: Primary Analysis**

| **Covariate** | **Coefficient (95% CI)** | **Standard error** | ***P* value** | **IRR (95% CI)** |
| --- | --- | --- | --- | --- |
| Active GLP-1RA prescription | −0.050 (−0.089, −0.011) | 0.020 | 0.011 | 0.951 (0.915, 0.989) |
| **Age** |  |  |  |  |
| <40 | 0.335 (0.264, 0.407) | 0.036 | <0.001 | 1.399 (1.303, 1.502) |
| 40-49 | 0.179 (0.116, 0.241) | 0.032 | <0.001 | 1.195 (1.123, 1.273) |
| 50-59 | 0.127 (0.073, 0.182) | 0.028 | <0.001 | 1.136 (1.075, 1.200) |
| 60-69 | - | - | - | - |
| 70-79 | −0.050 (−0.123, 0.024) | 0.037 | 0.184 | 0.952 (0.884, 1.024) |
| 80+ | 0.041 (−0.153, 0.231) | 0.098 | 0.673 | 1.042 (0.858, 1.260) |
| **Body mass index** |  |  |  |  |
| 18.5-24.9 | - | - | - | - |
| 25-29.9 | −0.089 (−0.206, 0.028) | 0.060 | 0.133 | 0.914 (0.814, 1.028) |
| 30-34.9 | −0.149 (−0.262, −0.035) | 0.058 | 0.010 | 0.862 (0.770, 0.966) |
| 35-39.9 | −0.176 (−0.290, −0.061) | 0.058 | 0.003 | 0.839 (0.748, 0.941) |
| 40+ | −0.222 (−0.336, −0.107) | 0.058 | <0.001 | 0.801 (0.715, 0.899) |
| **Charlson Comorbidity Index^a^** |  |  |  |  |
| 0 | - | - | - | - |
| 1-2 | −0.019 (−0.064, 0.026) | 0.023 | 0.406 | 0.981 (0.938, 1.026) |
| 3-4 | −0.020 (−0.086, 0.046) | 0.033 | 0.553 | 0.980 (0.918, 1.047) |
| 5+ | −0.117 (−0.203, −0.033) | 0.044 | 0.007 | 0.889 (0.817, 0.968) |
| **Health insurance** |  |  |  |  |
| Purchased directly/obtained through employer | - | - | - | - |
| Medicare/Medicaid dual enrollees | −0.327 (−0.427, −0.227) | 0.051 | <0.001 | 0.721 (0.652, 0.797) |
| Medicaid | −0.082 (−0.146, −0.018) | 0.033 | 0.012 | 0.921 (0.864, 0.982) |
| Medicare | −0.109 (−0.176, −0.042) | 0.034 | 0.001 | 0.897 (0.839, 0.958) |
| None | −0.036 (−0.164, 0.090) | 0.065 | 0.575 | 0.964 (0.849, 1.094) |
| Other | −0.117 (−0.219, −0.017) | 0.052 | 0.023 | 0.889 (0.804, 0.983) |
| Skip/prefer not to answer | −0.042 (−0.109, 0.025) | 0.034 | 0.221 | 0.959 (0.897, 1.025) |
| **Highest level of education** |  |  |  |  |
| Advanced degree | −0.012 (−0.070, 0.046) | 0.029 | 0.681 | 0.988 (0.932, 1.047) |
| College graduate | −0.025 (−0.078, 0.029) | 0.027 | 0.365 | 0.976 (0.925, 1.029) |
| Some college | - | - | - | - |
| Less than high school | 0.233 (0.149, 0.318) | 0.043 | <0.001 | 1.263 (1.160, 1.374) |
| Prefer not to answer | −0.034 (−0.210, 0.140) | 0.089 | 0.704 | 0.967 (0.810, 1.150) |
| High school or equivalent | 0.067 (0.008, 0.125) | 0.030 | 0.026 | 1.069 (1.008, 1.134) |
| **Household income** |  |  |  |  |
| <$50 000 | - | - | - | - |
| $50 000-$100 000 | 0.104 (0.049, 0.160) | 0.029 | <0.001 | 1.110 (1.050, 1.174) |
| $100 000+ | 0.216 (0.155, 0.278) | 0.032 | <0.001 | 1.242 (1.167, 1.321) |
| Prefer not to answer/skipped | 0.030 (−0.032, 0.093) | 0.032 | 0.341 | 1.031 (0.968, 1.097) |
| **Health history** |  |  |  |  |
| Current or former tobacco use | 0.171 (0.127, 0.215) | 0.022 | <0.001 | 1.186 (1.135, 1.239) |
| History of substance use | 0.213 (0.170, 0.255) | 0.022 | <0.001 | 1.237 (1.186, 1.291) |
| Depression | −0.011 (−0.053, 0.030) | 0.021 | 0.597 | 0.989 (0.948, 1.031) |
| Liver disease | 0.007 (−0.042, 0.056) | 0.025 | 0.765 | 1.008 (0.959, 1.058) |
| Hypertension | −0.063 (−0.123, −0.003) | 0.031 | 0.039 | 0.939 (0.884, 0.997) |
| Type 1 diabetes | −0.211 (−0.286, −0.137) | 0.038 | <0.001 | 0.810 (0.751, 0.872) |
| Type 2 diabetes | −0.167 (−0.216, −0.119) | 0.025 | <0.001 | 0.846 (0.806, 0.888) |
| Any off-label non-GLP-1RA AUD medication^b^ | −0.062 (−0.109, −0.016) | 0.024 | 0.009 | 0.940 (0.897, 0.984) |
| Any non-GLP-1RA obesity medication^c^ | −0.017 (−0.058, 0.023) | 0.021 | 0.403 | 0.983 (0.944, 1.024) |
| **Race/ethnicity** |  |  |  |  |
| Non-Hispanic (NH) White | - | - | - | - |
| NH Asian | −0.204 (−0.369, −0.042) | 0.084 | 0.015 | 0.815 (0.691, 0.959) |
| NH Black or African American | 0.069 (0.014, 0.123) | 0.028 | 0.013 | 1.071 (1.014, 1.131) |
| Hispanic | 0.007 (−0.055, 0.070) | 0.032 | 0.815 | 1.007 (0.947, 1.072) |
| NH more than one population | −0.017 (−0.110, 0.076) | 0.047 | 0.725 | 0.984 (0.896, 1.079) |
| NH other/none/unknown | −0.093 (−0.204, 0.016) | 0.056 | 0.096 | 0.911 (0.816, 1.016) |
| **Sex** |  |  |  |  |
| Female | - | - | - | - |
| Male | 0.231 (0.188, 0.274) | 0.022 | <0.001 | 1.260 (1.207, 1.315) |
| Unknown/other | 0.406 (0.197, 0.611) | 0.105 | <0.001 | 1.500 (1.218, 1.843) |
| **Visit days** |  |  |  |  |
| 1-2 | −0.104 (−0.186, −0.022) | 0.042 | 0.013 | 0.901 (0.830, 0.978) |
| 3-5 | 0.085 (−0.002, 0.172) | 0.044 | 0.054 | 1.089 (0.998, 1.187) |
| 6-10 | 0.041 (−0.029, 0.111) | 0.036 | 0.249 | 1.042 (0.971, 1.118) |
| 11-30 | 0.091 (0.046, 0.137) | 0.023 | <0.001 | 1.096 (1.047, 1.147) |
| 31+ | - | - | - | - |

Abbreviations: AUD, alcohol use disorder; CI, confidence interval; GLP-1RA, glucagon-like peptide-1 receptor agonist; IRR, incidence rate ratio.

^a^ Excludes liver disease and type 2 diabetes.

^b^ Phentermine, phendimetrazine, diethylpropion, phentermine-topiramate, naltrexone-bupropion, orlistat, metformin, topiramate, bupropion.

^c^ Topiramate, gabapentin, baclofen, varenicline, prazosin, doxazosin.

### **eTable 7. Negative Binomial Model Evaluating Association of Alcohol Use (AUDIT-C) with Active GLP-1RA Prescriptions Compared to Future GLP-1RA Prescriptions: Female Sex Primary Analysis**

| **Covariate** | **Coefficient (95% CI)** | **Standard error** | ***P* value** | **IRR (95% CI)** |
| --- | --- | --- | --- | --- |
| Active GLP-1RA prescription | −0.049 (−0.097, −0.002) | 0.024 | 0.043 | 0.952 (0.908, 0.998) |
| **Age** | | | | |
| <40 | 0.392 (0.310, 0.473) | 0.042 | <0.001 | 1.479 (1.363, 1.605) |
| 40-49 | 0.234 (0.159, 0.308) | 0.038 | <0.001 | 1.263 (1.173, 1.361) |
| 50-59 | 0.174 (0.107, 0.241) | 0.034 | <0.001 | 1.190 (1.113, 1.273) |
| 60-69 | - | - | - | - |
| 70-79 | −0.012 (−0.119, 0.095) | 0.054 | 0.827 | 0.988 (0.887, 1.099) |
| 80+ | 0.046 (−0.302, 0.374) | 0.172 | 0.788 | 1.047 (0.740, 1.453) |
| **Body mass index** | | | | |
| 18.5-24.9 | - | - | - | - |
| 25-29.9 | −0.055 (−0.202, 0.095) | 0.076 | 0.469 | 0.947 (0.817, 1.100) |
| 30-34.9 | −0.150 (−0.293, −0.005) | 0.073 | 0.040 | 0.860 (0.746, 0.995) |
| 35-39.9 | −0.152 (−0.296, −0.006) | 0.074 | 0.040 | 0.859 (0.744, 0.994) |
| 40+ | −0.229 (−0.372, −0.084) | 0.073 | 0.002 | 0.795 (0.689, 0.920) |
| **Charlson Comorbidity Index^a^** | | | | |
| 0 | - | - | - | - |
| 1-2 | −0.033 (−0.086, 0.019) | 0.027 | 0.215 | 0.967 (0.917, 1.020) |
| 3-4 | −0.098 (−0.182, −0.015) | 0.043 | 0.021 | 0.907 (0.834, 0.985) |
| 5+ | −0.219 (−0.335, −0.104) | 0.059 | <0.001 | 0.803 (0.715, 0.901) |
| **Health insurance** | | | | |
| Purchased directly/obtained through employer | - | - | - | - |
| Medicare/Medicaid dual enrollees | −0.277 (−0.394, −0.162) | 0.059 | <0.001 | 0.758 (0.674, 0.851) |
| Medicaid | −0.083 (−0.157, −0.009) | 0.038 | 0.028 | 0.920 (0.855, 0.991) |
| Medicare | −0.067 (−0.156, 0.021) | 0.045 | 0.134 | 0.935 (0.856, 1.021) |
| None | 0.052 (−0.101, 0.203) | 0.078 | 0.504 | 1.053 (0.904, 1.225) |
| Other | −0.057 (−0.186, 0.070) | 0.065 | 0.383 | 0.945 (0.830, 1.072) |
| Skip/prefer not to answer | −0.022 (−0.101, 0.057) | 0.040 | 0.588 | 0.978 (0.904, 1.058) |
| **Highest level of education** | | | | |
| Advanced degree | 0.015 (−0.055, 0.085) | 0.036 | 0.672 | 1.015 (0.947, 1.088) |
| College graduate | 0.005 (−0.059, 0.068) | 0.032 | 0.886 | 1.005 (0.943, 1.071) |
| Some college | - | - | - | - |
| Less than high school | 0.264 (0.162, 0.364) | 0.052 | <0.001 | 1.302 (1.176, 1.440) |
| Prefer not to answer | −0.199 (−0.436, 0.030) | 0.119 | 0.095 | 0.820 (0.646, 1.030) |
| High school or equivalent | 0.073 (0.002, 0.144) | 0.036 | 0.042 | 1.076 (1.002, 1.155) |
| **Household income** | | | | |
| <$50 000 | - | - | - | - |
| $50 000-$100 000 | 0.078 (0.010, 0.145) | 0.034 | 0.024 | 1.081 (1.011, 1.156) |
| $100 000+ | 0.203 (0.128, 0.279) | 0.038 | <0.001 | 1.225 (1.136, 1.321) |
| Prefer not to answer/skipped | 0.036 (−0.039, 0.111) | 0.038 | 0.345 | 1.037 (0.962, 1.117) |
| **Health history** | | | | |
| Current or former tobacco use | 0.146 (0.096, 0.197) | 0.026 | <0.001 | 1.158 (1.101, 1.217) |
| History of substance use | 0.231 (0.180, 0.282) | 0.026 | <0.001 | 1.260 (1.197, 1.326) |
| Depression | 0.033 (−0.017, 0.083) | 0.025 | 0.194 | 1.034 (0.983, 1.086) |
| Liver disease | 0.005 (−0.055, 0.065) | 0.031 | 0.877 | 1.005 (0.946, 1.067) |
| Hypertension | −0.069 (−0.133, −0.004) | 0.033 | 0.037 | 0.934 (0.876, 0.996) |
| Type 1 diabetes | −0.190 (−0.284, −0.098) | 0.048 | <0.001 | 0.827 (0.753, 0.907) |
| Type 2 diabetes | −0.162 (−0.218, −0.106) | 0.028 | <0.001 | 0.850 (0.805, 0.899) |
| Any off-label non-GLP-1RA AUD medication^b^ | −0.021 (−0.076, 0.035) | 0.028 | 0.468 | 0.980 (0.927, 1.036) |
| Any non-GLP-1RA obesity medication^c^ | −0.014 (−0.062, 0.035) | 0.025 | 0.582 | 0.986 (0.940, 1.036) |
| **Race/ethnicity** | | | | |
| Non-Hispanic (NH) White | - | - | - | - |
| NH Asian | −0.452 (−0.693, −0.221) | 0.120 | <0.001 | 0.636 (0.500, 0.802) |
| NH Black or African American | 0.087 (0.025, 0.149) | 0.032 | 0.006 | 1.091 (1.025, 1.161) |
| Hispanic | −0.057 (−0.131, 0.017) | 0.038 | 0.131 | 0.945 (0.878, 1.017) |
| NH more than one population | 0.003 (−0.105, 0.109) | 0.054 | 0.963 | 1.003 (0.900, 1.115) |
| NH other/none/unknown | −0.174 (−0.322, −0.030) | 0.074 | 0.019 | 0.840 (0.725, 0.971) |
| **Visit Days** | | | | |
| 1-2 | −0.124 (−0.222, −0.027) | 0.050 | 0.012 | 0.883 (0.801, 0.973) |
| 3-5 | 0.093 (−0.009, 0.194) | 0.052 | 0.074 | 1.097 (0.991, 1.215) |
| 6-10 | 0.055 (−0.027, 0.137) | 0.042 | 0.186 | 1.057 (0.974, 1.147) |
| 11-30 | 0.071 (0.016, 0.125) | 0.028 | 0.011 | 1.073 (1.016, 1.133) |
| 31+ | - | - | - | - |

Abbreviations: AUD, alcohol use disorder; CI, confidence interval; GLP-1RA, glucagon-like peptide-1 receptor agonist; IRR, incidence rate ratio.

^a^ Excludes liver disease and type 2 diabetes.

^b^ Phentermine, phendimetrazine, diethylpropion, phentermine-topiramate, naltrexone-bupropion, orlistat, metformin, topiramate, bupropion.

^c^ Topiramate, gabapentin, baclofen, varenicline, prazosin, doxazosin.

### **eTable 8. Negative Binomial Model Evaluating Association of Alcohol Use (AUDIT-C) with Active GLP-1RA Prescriptions Compared to Future GLP-1RA Prescriptions: Male Sex Primary Analysis**

| **Covariate** | **Coefficient (95% CI)** | **Standard error** | ***P* value** | **IRR (95% CI)** |
| --- | --- | --- | --- | --- |
| Active GLP-1RA prescription | −0.051 (−0.119, 0.018) | 0.035 | 0.146 | 0.951 (0.888, 1.018) |
| **Age** | | | | |
| <40 | 0.194 (0.047, 0.341) | 0.075 | 0.010 | 1.214 (1.048, 1.406) |
| 40-49 | 0.081 (−0.038, 0.199) | 0.061 | 0.184 | 1.084 (0.963, 1.221) |
| 50-59 | 0.059 (−0.038, 0.155) | 0.049 | 0.232 | 1.060 (0.963, 1.168) |
| 60-69 | - | - | - | - |
| 70-79 | −0.117 (−0.226, −0.009) | 0.055 | 0.032 | 0.889 (0.798, 0.991) |
| 80+ | 0.007 (−0.245, 0.258) | 0.127 | 0.953 | 1.008 (0.782, 1.294) |
| **Body mass index** | | | | |
| 18.5-24.9 | - | - | - | - |
| 25-29.9 | −0.118 (−0.315, 0.078) | 0.100 | 0.236 | 0.889 (0.730, 1.082) |
| 30-34.9 | −0.140 (−0.331, 0.052) | 0.097 | 0.150 | 0.870 (0.718, 1.053) |
| 35-39.9 | −0.255 (−0.451, −0.059) | 0.100 | 0.010 | 0.775 (0.637, 0.943) |
| 40+ | −0.222 (−0.419, −0.025) | 0.100 | 0.027 | 0.801 (0.658, 0.975) |
| **Charlson Comorbidity Index^a^** | | | | |
| 0 | - | - | - | - |
| 1-2 | −0.001 (−0.088, 0.085) | 0.044 | 0.974 | 0.999 (0.916, 1.089) |
| 3-4 | 0.078 (−0.033, 0.190) | 0.057 | 0.166 | 1.081 (0.968, 1.209) |
| 5+ | −0.011 (−0.147, 0.125) | 0.070 | 0.878 | 0.989 (0.864, 1.133) |
| **Health insurance** | | | | |
| Purchased directly/obtained through employer | - | - | - | - |
| Medicare/Medicaid dual enrollees | −0.461 (−0.656, −0.269) | 0.098 | <0.001 | 0.630 (0.519, 0.764) |
| Medicaid | −0.060 (−0.185, 0.064) | 0.063 | 0.339 | 0.941 (0.831, 1.066) |
| Medicare | −0.172 (−0.280, −0.064) | 0.054 | 0.002 | 0.842 (0.756, 0.938) |
| None | −0.235 (−0.464, −0.009) | 0.116 | 0.042 | 0.790 (0.629, 0.992) |
| Other | −0.153 (−0.319, 0.012) | 0.085 | 0.070 | 0.858 (0.727, 1.012) |
| Skip/prefer not to answer | −0.125 (−0.252, 0.002) | 0.064 | 0.052 | 0.882 (0.777, 1.002) |
| **Highest level of education** | | | | |
| Advanced degree | −0.073 (−0.175, 0.030) | 0.052 | 0.164 | 0.930 (0.839, 1.030) |
| College graduate | −0.080 (−0.178, 0.017) | 0.050 | 0.106 | 0.923 (0.837, 1.017) |
| Some college | - | - | - | - |
| Less than high school | 0.178 (0.020, 0.336) | 0.080 | 0.027 | 1.195 (1.020, 1.400) |
| Prefer not to answer | 0.036 (−0.288, 0.356) | 0.163 | 0.827 | 1.036 (0.749, 1.428) |
| High school or equivalent | 0.016 (−0.090, 0.122) | 0.053 | 0.764 | 1.016 (0.914, 1.130) |
| **Household income** | | | | |
| <$50 000 | - | - | - | - |
| $50 000-$100 000 | 0.133 (0.034, 0.233) | 0.051 | 0.009 | 1.143 (1.034, 1.262) |
| $100 000+ | 0.222 (0.114, 0.331) | 0.056 | <0.001 | 1.249 (1.121, 1.392) |
| Prefer not to answer/skipped | −0.015 (−0.129, 0.100) | 0.058 | 0.800 | 0.985 (0.879, 1.105) |
| **Health history** | | | | |
| Current or former tobacco use | 0.197 ( 0.111, 0.283) | 0.044 | <0.001 | 1.218 (1.117, 1.327) |
| History of substance use | 0.221 ( 0.146, 0.296) | 0.038 | <0.001 | 1.247 (1.157, 1.344) |
| Depression | −0.068 (−0.144, 0.009) | 0.039 | 0.080 | 0.935 (0.866, 1.009) |
| Liver disease | 0.001 (−0.086, 0.088) | 0.044 | 0.977 | 1.001 (0.918, 1.092) |
| Hypertension | −0.024 (−0.163, 0.115) | 0.071 | 0.735 | 0.976 (0.850, 1.121) |
| Type 1 diabetes | −0.251 (−0.382, −0.119) | 0.068 | <0.001 | 0.778 (0.683, 0.888) |
| Type 2 diabetes | −0.179 (−0.275, −0.083) | 0.049 | <0.001 | 0.836 (0.759, 0.921) |
| Any off-label non-GLP-1RA AUD medication^b^ | −0.164 (−0.247, −0.080) | 0.042 | <0.001 | 0.849 (0.781, 0.923) |
| Any non-GLP-1RA obesity medication^c^ | −0.027 (−0.101, 0.047) | 0.038 | 0.468 | 0.973 (0.904, 1.048) |
| **Race/ethnicity** | | | | |
| Non-Hispanic (NH) White | - | - | - | - |
| NH Asian | −0.204 (−0.369, −0.042) | 0.084 | 0.015 | 0.815 (0.691, 0.959) |
| NH Black or African American | 0.069 (0.014, 0.123) | 0.028 | 0.013 | 1.071 (1.014, 1.131) |
| Hispanic | 0.007 (−0.055, 0.070) | 0.032 | 0.815 | 1.007 (0.947, 1.072) |
| NH more than one population | −0.017 (−0.110, 0.076) | 0.047 | 0.725 | 0.984 (0.896, 1.079) |
| NH other/none/unknown | −0.093 (−0.204, 0.016) | 0.056 | 0.096 | 0.911 (0.816, 1.016) |
| **Visit days** | | | | |
| 1-2 | −0.028 (−0.182, 0.125) | 0.078 | 0.718 | 0.972 (0.833, 1.134) |
| 3-5 | 0.037 (−0.126, 0.199) | 0.083 | 0.658 | 1.037 (0.882, 1.220) |
| 6-10 | 0.004 (−0.129, 0.136) | 0.067 | 0.955 | 1.004 (0.879, 1.146) |
| 11-30 | 0.137 (0.053, 0.221) | 0.043 | 0.001 | 1.147 (1.054, 1.248) |
| 31+ | - | - | - | - |

Abbreviations: AUD, alcohol use disorder; CI, confidence interval; GLP-1RA, glucagon-like peptide-1 receptor agonist; IRR, incidence rate ratio.

^a^ Excludes liver disease and type 2 diabetes.

^b^ Phentermine, phendimetrazine, diethylpropion, phentermine-topiramate, naltrexone-bupropion, orlistat, metformin, topiramate, bupropion.

^c^ Topiramate, gabapentin, baclofen, varenicline, prazosin, doxazosin.

### **eTable 9. Negative Binomial Model Evaluating Association of Alcohol Use (AUDIT-C) with Former GLP-1RA Prescriptions Compared to Future GLP-1RA Prescriptions: Primary Analysis**

| **Covariate** | **Coefficient (95% CI)** | **Standard error** | ***P* value** | **IRR (95% CI)** |
| --- | --- | --- | --- | --- |
| Former GLP-1RA prescription | −0.079 (−0.166, 0.007) | 0.044 | 0.073 | 0.924 (0.847, 1.007) |
| **Age** | | | | |
| <40 | 0.365 (0.282, 0.448) | 0.042 | <0.001 | 1.440 (1.326, 1.564) |
| 40-49 | 0.190 (0.114, 0.266) | 0.039 | <0.001 | 1.209 (1.120, 1.305) |
| 50-59 | 0.159 (0.092, 0.227) | 0.034 | <0.001 | 1.173 (1.096, 1.255) |
| 60-69 | - | - | - | - |
| 70-79 | −0.049 (−0.143, 0.044) | 0.047 | 0.301 | 0.952 (0.867, 1.045) |
| 80+ | 0.008 (−0.246, 0.256) | 0.128 | 0.949 | 1.008 (0.782, 1.292) |
| **Body mass index** | | | | |
| 18.5-24.9 | - | - | - | - |
| 25-29.9 | −0.083 (−0.225, 0.059) | 0.072 | 0.247 | 0.920 (0.799, 1.061) |
| 30-34.9 | −0.136 (−0.272, 0.001) | 0.070 | 0.051 | 0.873 (0.762, 1.001) |
| 35-39.9 | −0.167 (−0.305, −0.028) | 0.070 | 0.017 | 0.846 (0.737, 0.972) |
| 40+ | −0.253 (−0.391, −0.115) | 0.070 | <0.001 | 0.776 (0.676, 0.892) |
| **Charlson Comorbidity Index^a^** | | | | |
| 0 | - | - | - | - |
| 1-2 | 0.010 (−0.045, 0.065) | 0.028 | 0.713 | 1.010 (0.956, 1.067) |
| 3-4 | 0.024 (−0.057, 0.104) | 0.041 | 0.564 | 1.024 (0.945, 1.109) |
| 5+ | −0.048 (−0.154, 0.057) | 0.054 | 0.370 | 0.953 (0.857, 1.058) |
| **Health insurance** | | | | |
| Purchased directly/obtained through employer | - | - | - | - |
| Medicare/Medicaid dual enrollees | −0.293 (−0.418, −0.168) | 0.064 | <0.001 | 0.746 (0.658, 0.845) |
| Medicaid | −0.097 (−0.174, −0.021) | 0.039 | 0.013 | 0.907 (0.840, 0.979) |
| Medicare | −0.057 (−0.140, 0.025) | 0.042 | 0.175 | 0.945 (0.870, 1.026) |
| None | −0.055 (−0.208, 0.096) | 0.077 | 0.475 | 0.946 (0.812, 1.100) |
| Other/prefer not to answer | −0.074 (−0.145, −0.004) | 0.036 | 0.039 | 0.928 (0.865, 0.996) |
| **Highest level of education** | | | | |
| Advanced degree | 0.037 (−0.034, 0.109) | 0.036 | 0.305 | 1.038 (0.967, 1.115) |
| College graduate | 0.011 (−0.054, 0.077) | 0.034 | 0.732 | 1.012 (0.947, 1.080) |
| Some college | - | - | - | - |
| Less than high school | 0.217 (0.115, 0.319) | 0.052 | <0.001 | 1.242 (1.122, 1.375) |
| Prefer not to answer | 0.037 (−0.164, 0.235) | 0.101 | 0.716 | 1.038 (0.848, 1.265) |
| High school or equivalent | 0.119 (0.048, 0.190) | 0.036 | 0.001 | 1.126 (1.049, 1.209) |
| **Household income** | | | | |
| <$50 000 | - | - | - | - |
| $50 000-$100 000 | 0.084 (0.015, 0.153) | 0.035 | 0.017 | 1.088 (1.015, 1.165) |
| $100 000+ | 0.211 (0.135, 0.287) | 0.039 | <0.001 | 1.235 (1.145, 1.333) |
| Prefer not to answer/skipped | −0.001 (−0.078, 0.076) | 0.039 | 0.981 | 0.999 (0.925, 1.078) |
| **Health history** | | | | |
| Current or former tobacco use | 0.175 (0.123, 0.228) | 0.027 | <0.001 | 1.192 (1.131, 1.256) |
| History of substance use | 0.228 (0.176, 0.279) | 0.026 | <0.001 | 1.256 (1.193, 1.322) |
| Depression | −0.014 (−0.065, 0.038) | 0.026 | 0.599 | 0.986 (0.937, 1.038) |
| Liver disease | −0.005 (−0.068, 0.058) | 0.032 | 0.869 | 0.995 (0.934, 1.059) |
| Hypertension | 0.016 (−0.054, 0.086) | 0.036 | 0.657 | 1.016 (0.947, 1.090) |
| Type 1 diabetes | −0.215 (−0.312, −0.117) | 0.050 | <0.001 | 0.807 (0.732, 0.889) |
| Type 2 diabetes | −0.152 (−0.209, −0.095) | 0.029 | <0.001 | 0.859 (0.811, 0.909) |
| Any off-label non-GLP-1RA AUD medication^b^ | −0.085 (−0.143, −0.027) | 0.030 | 0.004 | 0.919 (0.866, 0.974) |
| Any non-GLP-1RA obesity medication^c^ | −0.015 (−0.065, 0.036) | 0.026 | 0.568 | 0.985 (0.937, 1.036) |
| **Race/ethnicity** | | | | |
| Non-Hispanic (NH) White | - | - | - | - |
| NH Asian | −0.265 (−0.479, −0.056) | 0.108 | 0.014 | 0.767 (0.619, 0.946) |
| NH Black or African American | 0.067 (0.001, 0.133) | 0.033 | 0.044 | 1.069 (1.001, 1.142) |
| Hispanic | 0.020 (−0.056, 0.097) | 0.039 | 0.604 | 1.020 (0.945, 1.101) |
| NH more than one population | 0.001 (−0.113, 0.115) | 0.058 | 0.983 | 1.001 (0.893, 1.122) |
| NH other/none/unknown | −0.135 (−0.271, <0.001) | 0.069 | 0.052 | 0.874 (0.763, 1.000) |
| **Sex** | | | | |
| Female | - | - | - | - |
| Male | 0.215 (0.162, 0.267) | 0.027 | <0.001 | 1.239 (1.176, 1.306) |
| Unknown/other | 0.368 (0.119, 0.613) | 0.125 | 0.003 | 1.445 (1.127, 1.846) |
| **Visit days** | | | | |
| 1-2 | −0.044 (−0.145, 0.055) | 0.051 | 0.384 | 0.957 (0.865, 1.057) |
| 3-5 | 0.077 (−0.023, 0.176) | 0.051 | 0.130 | 1.080 (0.978, 1.193) |
| 6-10 | 0.041 (−0.042, 0.123) | 0.042 | 0.334 | 1.042 (0.959, 1.131) |
| 11-30 | 0.085 (0.029, 0.142) | 0.029 | 0.003 | 1.089 (1.030, 1.152) |
| 31+ | - | - | - | - |

Abbreviations: AUD, alcohol use disorder; CI, confidence interval; GLP-1RA, glucagon-like peptide-1 receptor agonist; IRR, incidence rate ratio.

^a^ Excludes liver disease and type 2 diabetes.

^b^ Phentermine, phendimetrazine, diethylpropion, phentermine-topiramate, naltrexone-bupropion, orlistat, metformin, topiramate, bupropion.

^c^ Topiramate, gabapentin, baclofen, varenicline, prazosin, doxazosin.

### **eTable 10. Negative Binomial Model Evaluating Association of Alcohol Use (AUDIT-C) with Active GLP-1RA Prescriptions Compared to Matched Group: Secondary Analysis**

| **Covariate** | **Coefficient (95% CI)** | **Standard error** | ***P* value** | **IRR (95% CI)** |
| --- | --- | --- | --- | --- |
| Active GLP-1RA prescription | −0.114 (−0.159, −0.069) | 0.023 | <0.001 | 0.892 (0.853, 0.933) |
| **Age** | | | | |
| <40 | 0.339 (0.244, 0.433) | 0.048 | <0.001 | 1.403 (1.277, 1.541) |
| 40-49 | 0.221 (0.147, 0.296) | 0.038 | <0.001 | 1.248 (1.158, 1.344) |
| 50-59 | 0.119 (0.057, 0.182) | 0.032 | <0.001 | 1.127 (1.058, 1.200) |
| 60-69 | - | - | - | - |
| 70-79 | −0.034 (−0.110, 0.043) | 0.039 | 0.391 | 0.967 (0.895, 1.044) |
| 80+ | −0.107 (−0.291, 0.074) | 0.093 | 0.251 | 0.899 (0.747, 1.077) |
| **Body mass index** | | | | |
| 18.5-24.9 | - | - | - | - |
| 25-29.9 | 0.071 (−0.061, 0.203) | 0.067 | 0.295 | 1.073 (0.941, 1.225) |
| 30-34.9 | −0.003 (−0.132, 0.126) | 0.066 | 0.959 | 0.997 (0.876, 1.134) |
| 35-39.9 | −0.071 (−0.201, 0.061) | 0.067 | 0.292 | 0.932 (0.818, 1.063) |
| 40+ | −0.121 (−0.251, 0.011) | 0.067 | 0.071 | 0.886 (0.778, 1.011) |
| **Charlson Comorbidity Index^a^** | | | | |
| 0 | - | - | - | - |
| 1-2 | −0.059 (−0.113, −0.005) | 0.028 | 0.033 | 0.943 (0.893, 0.995) |
| 3-4 | −0.064 (−0.140, 0.012) | 0.039 | 0.098 | 0.938 (0.870, 1.012) |
| 5+ | −0.147 (−0.239, −0.055) | 0.047 | 0.002 | 0.864 (0.788, 0.947) |
| **Health insurance** | | | | |
| Purchased directly/obtained through employer | - | - | - | - |
| Medicare/Medicaid dual enrollees | −0.290 (−0.400, −0.180) | 0.056 | <0.001 | 0.748 (0.670, 0.835) |
| Medicaid | −0.056 (−0.134, 0.023) | 0.040 | 0.163 | 0.946 (0.875, 1.023) |
| Medicare | −0.095 (−0.168, −0.023) | 0.037 | 0.010 | 0.909 (0.846, 0.977) |
| None | 0.065 (−0.087, 0.217) | 0.078 | 0.399 | 1.068 (0.916, 1.243) |
| Other | −0.118 (−0.229, −0.007) | 0.057 | 0.039 | 0.889 (0.795, 0.993) |
| Skip/prefer not to answer | 0.012 (−0.090, 0.113) | 0.052 | 0.823 | 1.012 (0.914, 1.119) |
| **Highest level of education** | | | | |
| Advanced degree | −0.014 (−0.081, 0.053) | 0.034 | 0.681 | 0.986 (0.922, 1.055) |
| College graduate | −0.011 (−0.073, 0.051) | 0.032 | 0.727 | 0.989 (0.930, 1.052) |
| Some college | - | - | - | - |
| Less than high school | 0.071 (−0.040, 0.181) | 0.057 | 0.212 | 1.073 (0.961, 1.198) |
| Prefer not to answer | 0.197 (−0.024, 0.416) | 0.112 | 0.078 | 1.218 (0.977, 1.516) |
| High school or equivalent | 0.069 (−0.001, 0.139) | 0.036 | 0.051 | 1.072 (0.999, 1.150) |
| **Household income** | | | | |
| <$50 000 | - | - | - | - |
| $50 000-$100 000 | 0.136 (0.071, 0.200) | 0.033 | <0.001 | 1.145 (1.073, 1.222) |
| $100 000+ | 0.236 (0.163, 0.309) | 0.037 | <0.001 | 1.266 (1.178, 1.362) |
| Prefer not to answer/skipped | 0.009 (−0.065, 0.083) | 0.038 | 0.809 | 1.009 (0.937, 1.087) |
| **Health history** | | | | |
| Current or former tobacco use | 0.230 (0.177, 0.283) | 0.027 | <0.001 | 1.259 (1.194, 1.328) |
| History of substance use | 0.228 (0.177, 0.278) | 0.026 | <0.001 | 1.256 (1.194, 1.320) |
| Depression | −0.050 (−0.099, −0.001) | 0.025 | 0.046 | 0.951 (0.906, 0.999) |
| Liver disease | 0.014 (−0.040, 0.069) | 0.028 | 0.604 | 1.015 (0.961, 1.071) |
| Hypertension | −0.014 (−0.098, 0.070) | 0.043 | 0.747 | 0.986 (0.907, 1.073) |
| Type 1 diabetes | −0.223 (−0.311, −0.136) | 0.045 | <0.001 | 0.800 (0.733, 0.873) |
| Type 2 diabetes | −0.182 (−0.250, −0.113) | 0.035 | <0.001 | 0.834 (0.779, 0.893) |
| Any off-label non-GLP-1RA AUD medication^b^ | −0.050 (−0.102, 0.002) | 0.026 | 0.058 | 0.951 (0.903, 1.002) |
| Any non-GLP-1RA obesity medication^c^ | 0.014 (−0.035, 0.063) | 0.025 | 0.576 | 1.014 (0.966, 1.065) |
| **Race/ethnicity** | | | | |
| Non-Hispanic (NH) White | - | - | - | - |
| NH Asian | −0.256 (−0.458, −0.057) | 0.102 | 0.012 | 0.774 (0.633, 0.944) |
| NH Black or African American | 0.154 (0.088, 0.220) | 0.033 | <0.001 | 1.166 (1.092, 1.246) |
| Hispanic | 0.053 (−0.024, 0.130) | 0.039 | 0.173 | 1.055 (0.977, 1.139) |
| NH more than one population | −0.060 (−0.172, 0.052) | 0.057 | 0.295 | 0.942 (0.842, 1.053) |
| NH other/none/unknown | −0.094 (−0.219, 0.029) | 0.063 | 0.137 | 0.910 (0.804, 1.030) |
| **Sex** | | | | |
| Female | - | - | - | - |
| Male | 0.233 (0.183, 0.282) | 0.025 | <0.001 | 1.262 (1.201, 1.326) |
| Unknown/other | 0.400 (0.171, 0.627) | 0.116 | 0.001 | 1.491 (1.186, 1.871) |
| **Visit days** | | | | |
| 1-2 | −0.154 (−0.246, −0.063) | 0.047 | 0.001 | 0.857 (0.782, 0.939) |
| 3-5 | 0.007 (−0.114, 0.128) | 0.062 | 0.908 | 1.007 (0.892, 1.137) |
| 6-10 | −0.011 (−0.104, 0.081) | 0.047 | 0.816 | 0.989 (0.902, 1.085) |
| 11-30 | 0.094 (0.039, 0.148) | 0.028 | 0.001 | 1.098 (1.040, 1.160) |
| 31+ | - | - | - | - |

Abbreviations: AUD, alcohol use disorder; CI, confidence interval; GLP-1RA, glucagon-like peptide-1 receptor agonist; IRR, incidence rate ratio.

^a^ Excludes liver disease and type 2 diabetes.

^b^ Phentermine, phendimetrazine, diethylpropion, phentermine-topiramate, naltrexone-bupropion, orlistat, metformin, topiramate, bupropion.

^c^ Topiramate, gabapentin, baclofen, varenicline, prazosin, doxazosin.

### **eTable 11. Negative Binomial Model Evaluating Association of Alcohol Use (AUDIT-C) with Former GLP-1RA Prescriptions Compared to Matched Group: Secondary Analysis**

| **Covariate** | **Coefficient (95% CI)** | **Standard error** | ***P* value** | **IRR (95% CI)** |
| --- | --- | --- | --- | --- |
| Former GLP-1RA prescription | −0.018 (−0.142, 0.105) | 0.062 | 0.768 | 0.982 (0.868, 1.111) |
| **Age** | | | | |
| <40 | 0.331 (0.085, 0.578) | 0.125 | 0.008 | 1.393 (1.089, 1.782) |
| 40-49 | 0.089 (−0.127, 0.306) | 0.111 | 0.419 | 1.094 (0.881, 1.358) |
| 50-59 | 0.012 (−0.162, 0.186) | 0.088 | 0.887 | 1.013 (0.851, 1.205) |
| 60-69 | - | - | - | - |
| 70-79 | −0.028 (−0.240, 0.184) | 0.107 | 0.796 | 0.973 (0.787, 1.202) |
| 80+ | −0.142 (−0.626, 0.327) | 0.239 | 0.553 | 0.868 (0.535, 1.387) |
| **Body mass index** | | | | |
| 18.5-24.9 | - | - | - | - |
| 25-29.9 | −0.022 (−0.320, 0.277) | 0.151 | 0.884 | 0.978 (0.726, 1.319) |
| 30-34.9 | −0.016 (−0.306, 0.276) | 0.148 | 0.916 | 0.984 (0.736, 1.318) |
| 35-39.9 | −0.076 (−0.373, 0.223) | 0.152 | 0.617 | 0.927 (0.688, 1.250) |
| 40+ | −0.140 (−0.437, 0.159) | 0.152 | 0.358 | 0.870 (0.646, 1.172) |
| **Charlson Comorbidity Index^a^** | | | | |
| 0 | - | - | - | - |
| 1-2 | 0.002 (−0.143, 0.147) | 0.074 | 0.983 | 1.002 (0.866, 1.158) |
| 3-4 | −0.107 (−0.333, 0.117) | 0.113 | 0.343 | 0.898 (0.717, 1.124) |
| 5+ | −0.148 (−0.412, 0.114) | 0.133 | 0.266 | 0.862 (0.662, 1.121) |
| **Health insurance** | | | | |
| Purchased directly/obtained through employer | - | - | - | - |
| Medicare/Medicaid dual enrollees | −0.399 (−0.730, −0.075) | 0.165 | 0.016 | 0.671 (0.482, 0.928) |
| Medicaid | 0.025 (−0.190, 0.241) | 0.109 | 0.816 | 1.026 (0.827, 1.273) |
| Medicare | −0.135 (−0.328, 0.057) | 0.098 | 0.167 | 0.874 (0.721, 1.059) |
| None | 0.139 (−0.370, 0.646) | 0.256 | 0.588 | 1.149 (0.691, 1.907) |
| Other/Prefer not to answer | 0.026 (−0.219, 0.271) | 0.124 | 0.833 | 1.027 (0.803, 1.311) |
| **Highest level of education** | | | | |
| Advanced degree | 0.018 (−0.171, 0.208) | 0.096 | 0.847 | 1.019 (0.843, 1.231) |
| College graduate | 0.072 (−0.104, 0.247) | 0.090 | 0.424 | 1.074 (0.901, 1.280) |
| Some college | - | - | - | - |
| Less than high school | 0.079 (−0.218, 0.375) | 0.149 | 0.595 | 1.083 (0.804, 1.455) |
| Prefer not to answer | −0.001 (−0.466, 0.456) | 0.233 | 0.998 | 0.999 (0.627, 1.578) |
| High school or equivalent | 0.114 (−0.084, 0.311) | 0.101 | 0.259 | 1.120 (0.919, 1.365) |
| **Household income** | | | | |
| <$50 000 | - | - | - | - |
| $50 000-$100 000 | 0.168 (−0.017, 0.353) | 0.094 | 0.075 | 1.183 (0.983, 1.423) |
| $100 000+ | 0.098 (−0.112, 0.307) | 0.107 | 0.362 | 1.102 (0.894, 1.359) |
| Prefer not to answer/skipped | −0.015 (−0.228, 0.197) | 0.108 | 0.888 | 0.985 (0.796, 1.217) |
| **Health history** | | | | |
| Current or former tobacco use | 0.199 (0.049, 0.350) | 0.076 | 0.009 | 1.221 (1.050, 1.419) |
| History of substance use | 0.298 (0.154, 0.441) | 0.072 | <0.001 | 1.347 (1.167, 1.555) |
| Depression | −0.068 (−0.204, 0.069) | 0.070 | 0.331 | 0.935 (0.816, 1.071) |
| Liver disease | 0.044 (−0.118, 0.206) | 0.082 | 0.590 | 1.045 (0.888, 1.229) |
| Hypertension | 0.073 (−0.158, 0.305) | 0.118 | 0.536 | 1.076 (0.854, 1.356) |
| Type 1 diabetes | −0.132 (−0.395, 0.131) | 0.132 | 0.320 | 0.877 (0.674, 1.140) |
| Type 2 diabetes | −0.181 (−0.352, −0.010) | 0.086 | 0.035 | 0.834 (0.703, 0.990) |
| Any off-label non-GLP-1RA AUD medication^b^ | −0.103 (−0.254, 0.047) | 0.076 | 0.172 | 0.902 (0.776, 1.048) |
| Any non-GLP-1RA obesity medication^c^ | 0.050 (−0.083, 0.183) | 0.068 | 0.463 | 1.051 (0.920, 1.201) |
| **Race/ethnicity** | | | | |
| Non-Hispanic (NH) White | - | - | - | - |
| NH Asian | −0.690 (-1.246, −0.172) | 0.269 | 0.010 | 0.502 (0.288, 0.842) |
| NH Black or African American | 0.080 (−0.120, 0.280) | 0.100 | 0.423 | 1.084 (0.887, 1.323) |
| Hispanic | −0.019 (−0.225, 0.186) | 0.105 | 0.853 | 0.981 (0.798, 1.204) |
| NH more than one population | −0.298 (−0.666, 0.059) | 0.185 | 0.106 | 0.742 (0.514, 1.061) |
| NH other/none/unknown | 0.052 (−0.293, 0.392) | 0.174 | 0.767 | 1.053 (0.746, 1.480) |
| **Sex** | | | | |
| Female | - | - | - | - |
| Male | 0.316 (0.175, 0.456) | 0.071 | <0.001 | 1.371 (1.191, 1.578) |
| Unknown/other | 0.445 (−0.201, 1.081) | 0.326 | 0.173 | 1.560 (0.818, 2.947) |
| **Visit Days** | | | | |
| 1-2 | −0.064 (−0.258, 0.130) | 0.099 | 0.520 | 0.938 (0.773, 1.138) |
| 3-5 | 0.003 (−0.311, 0.315) | 0.159 | 0.983 | 1.003 (0.732, 1.370) |
| 6-10 | 0.151 (−0.068, 0.369) | 0.112 | 0.177 | 1.163 (0.934, 1.447) |
| 11-30 | 0.078 (−0.081, 0.237) | 0.081 | 0.334 | 1.081 (0.922, 1.268) |
| 31+ | - | - | - | - |

Abbreviations: AUD, alcohol use disorder; CI, confidence interval; GLP-1RA, glucagon-like peptide-1 receptor agonist; IRR, incidence rate ratio.

^a^ Excludes liver disease and type 2 diabetes.

^b^ Phentermine, phendimetrazine, diethylpropion, phentermine-topiramate, naltrexone-bupropion, orlistat, metformin, topiramate, bupropion.

^c^ Topiramate, gabapentin, baclofen, varenicline, prazosin, doxazosin.

### **eTable 12. Negative Binomial Model Evaluating Association of Alcohol Use with Active GLP-1RA Prescriptions Compared to Future GLP-1RA Prescriptions: AUDIT-C Drink Frequency Question Analysis**

| **Covariate** | **Coefficient (95% CI)** | **Standard error** | ***P* value** | **IRR (95% CI)** |
| --- | --- | --- | --- | --- |
| Active GLP-1RA prescription | −0.046 (−0.083, −0.009) | 0.019 | 0.015 | 0.955 (0.920, 0.991) |
| **Age** | | | | |
| <40 | 0.100 (0.031, 0.168) | 0.035 | 0.004 | 1.105 (1.032, 1.183) |
| 40-49 | 0.029 (−0.031, 0.090) | 0.031 | 0.338 | 1.030 (0.970, 1.094) |
| 50-59 | 0.034 (−0.018, 0.086) | 0.027 | 0.204 | 1.034 (0.982, 1.090) |
| 60-69 | - | - | - | - |
| 70-79 | 0.046 (−0.021, 0.113) | 0.034 | 0.176 | 1.047 (0.979, 1.119) |
| 80+ | 0.095 (−0.085, 0.265) | 0.089 | 0.289 | 1.099 (0.919, 1.304) |
| **Body mass index** | | | | |
| 18.5-24.9 | - | - | - | - |
| 25-29.9 | −0.091 (−0.197, 0.018) | 0.055 | 0.099 | 0.913 (0.821, 1.019) |
| 30-34.9 | −0.117 (−0.220, −0.011) | 0.053 | 0.028 | 0.890 (0.803, 0.989) |
| 35-39.9 | −0.157 (−0.262, −0.049) | 0.054 | 0.004 | 0.855 (0.770, 0.952) |
| 40+ | −0.228 (−0.333, −0.120) | 0.054 | <0.001 | 0.796 (0.717, 0.887) |
| **Charlson Comorbidity Index^a^** | | | | |
| 0 | - | - | - | - |
| 1-2 | 0.001 (−0.042, 0.044) | 0.022 | 0.955 | 1.001 (0.959, 1.045) |
| 3-4 | −0.010 (−0.073, 0.052) | 0.032 | 0.755 | 0.990 (0.930, 1.054) |
| 5+ | −0.116 (−0.199, −0.033) | 0.042 | 0.006 | 0.891 (0.819, 0.967) |
| **Health insurance** | | | | |
| Purchased directly/obtained through employer | - | - | - | - |
| Medicare/Medicaid dual enrollees | −0.304 (−0.407, −0.203) | 0.052 | <0.001 | 0.738 (0.665, 0.816) |
| Medicaid | −0.086 (−0.150, −0.023) | 0.032 | 0.008 | 0.917 (0.861, 0.977) |
| Medicare | −0.063 (−0.126, −0.001) | 0.032 | 0.045 | 0.939 (0.882, 0.999) |
| None | −0.061 (−0.194, 0.067) | 0.066 | 0.357 | 0.941 (0.824, 1.069) |
| Other | −0.105 (−0.205, −0.008) | 0.050 | 0.035 | 0.900 (0.815, 0.992) |
| Skip/prefer not to answer | −0.049 (−0.113, 0.015) | 0.033 | 0.137 | 0.952 (0.893, 1.015) |
| **Highest level of education** | | | | |
| Advanced degree | 0.086 (0.033, 0.139) | 0.027 | 0.001 | 1.090 (1.033, 1.149) |
| College graduate | 0.038 (−0.012, 0.088) | 0.026 | 0.137 | 1.039 (0.988, 1.092) |
| Some college | - | - | - | - |
| Less than high school | −0.045 (−0.137, 0.046) | 0.047 | 0.336 | 0.956 (0.872, 1.047) |
| Prefer not to answer | −0.037 (−0.212, 0.131) | 0.087 | 0.676 | 0.964 (0.809, 1.140) |
| High school or equivalent | −0.067 (−0.127, −0.008) | 0.030 | 0.027 | 0.935 (0.881, 0.992) |
| **Household income** | | | | |
| <$50 000 | - | - | - | - |
| $50 000-$100 000 | 0.124 (0.071, 0.178) | 0.027 | <0.001 | 1.132 (1.073, 1.194) |
| $100 000+ | 0.259 (0.201, 0.317) | 0.030 | <0.001 | 1.296 (1.223, 1.373) |
| Prefer not to answer/skipped | 0.062 (<0.001, 0.124) | 0.031 | 0.049 | 1.064 (1.000, 1.131) |
| **Health history** | | | | |
| Current or former tobacco use | 0.126 (0.085, 0.168) | 0.021 | <0.001 | 1.135 (1.088, 1.183) |
| History of substance use | 0.135 (0.095, 0.176) | 0.021 | <0.001 | 1.145 (1.100, 1.192) |
| Depression | −0.052 (−0.091, −0.012) | 0.020 | 0.010 | 0.949 (0.913, 0.988) |
| Liver disease | −0.009 (−0.057, 0.038) | 0.024 | 0.707 | 0.991 (0.945, 1.039) |
| Hypertension | −0.034 (−0.090, 0.023) | 0.029 | 0.243 | 0.967 (0.914, 1.023) |
| Type 1 diabetes | −0.198 (−0.270, −0.126) | 0.037 | <0.001 | 0.820 (0.763, 0.882) |
| Type 2 diabetes | −0.160 (−0.206, −0.114) | 0.024 | <0.001 | 0.852 (0.814, 0.892) |
| Any off-label non-GLP-1RA AUD medication^b^ | −0.057 (−0.102, −0.012) | 0.023 | 0.014 | 0.945 (0.903, 0.988) |
| Any non-GLP-1RA obesity medication^c^ | −0.024 (−0.063, 0.015) | 0.020 | 0.233 | 0.977 (0.939, 1.015) |
| **Race/ethnicity** | | | | |
| Non-Hispanic (NH) White | - | - | - | - |
| NH Asian | −0.160 (−0.315, −0.011) | 0.077 | 0.039 | 0.852 (0.730, 0.989) |
| NH Black or African American | 0.035 (−0.018, 0.087) | 0.027 | 0.195 | 1.035 (0.982, 1.091) |
| Hispanic | −0.106 (−0.169, −0.043) | 0.032 | 0.001 | 0.900 (0.844, 0.958) |
| NH more than one population | −0.020 (−0.110, 0.067) | 0.045 | 0.656 | 0.980 (0.896, 1.070) |
| NH other/none/unknown | −0.126 (−0.236, −0.020) | 0.055 | 0.022 | 0.882 (0.790, 0.981) |
| **Sex** | | | | |
| Female | - | - | - | - |
| Male | 0.109 (0.068, 0.150) | 0.021 | <0.001 | 1.115 (1.070, 1.162) |
| Unknown/other | 0.301 (0.100, 0.491) | 0.100 | 0.003 | 1.352 (1.105, 1.634) |
| **Visit days** | | | | |
| 1-2 | −0.112 (−0.192, −0.033) | 0.040 | 0.006 | 0.894 (0.826, 0.967) |
| 3-5 | 0.043 (−0.041, 0.124) | 0.042 | 0.311 | 1.044 (0.960, 1.132) |
| 6-10 | −0.012 (−0.080, 0.054) | 0.034 | 0.716 | 0.988 (0.923, 1.056) |
| 11-30 | 0.067 (0.024, 0.111) | 0.022 | 0.002 | 1.070 (1.024, 1.117) |
| 31+ | - | - | - | - |

Abbreviations: AUD, alcohol use disorder; CI, confidence interval; GLP-1RA, glucagon-like peptide-1 receptor agonist; IRR, incidence rate ratio.

^a^ Excludes liver disease and type 2 diabetes.

^b^ Phentermine, phendimetrazine, diethylpropion, phentermine-topiramate, naltrexone-bupropion, orlistat, metformin, topiramate, bupropion.

^c^ Topiramate, gabapentin, baclofen, varenicline, prazosin, doxazosin.

### **eTable 13. Negative Binomial Model Evaluating Association of Alcohol Use with Active GLP-1RA Prescriptions Compared to Future GLP-1RA Prescriptions: AUDIT-C Drink Quantity Question Analysis**

| **Covariate** | **Coefficient (95% CI)** | **Standard error** | ***P* value** | **IRR (95% CI)** |
| --- | --- | --- | --- | --- |
| Active GLP-1RA prescription | −0.062 (−0.171, 0.047) | 0.055 | 0.264 | 0.940 (0.843, 1.048) |
| **Age** | | | | |
| <40 | 0.824 (0.634, 1.015) | 0.097 | <0.001 | 2.279 (1.885, 2.759) |
| 40-49 | 0.479 (0.302, 0.657) | 0.090 | <0.001 | 1.615 (1.353, 1.928) |
| 50-59 | 0.442 (0.285, 0.601) | 0.081 | <0.001 | 1.556 (1.329, 1.824) |
| 60-69 | - | - | - | - |
| 70-79 | −0.688 (−0.974, −0.413) | 0.144 | <0.001 | 0.503 (0.378, 0.662) |
| 80+ | −0.957 (-1.954, −0.157) | 0.469 | 0.041 | 0.384 (0.142, 0.855) |
| **Body mass index** | | | | |
| 18.5-24.9 | **-** | **-** | **-** | **-** |
| 25-29.9 | −0.219 (−0.537, 0.109) | 0.164 | 0.181 | 0.803 (0.584, 1.115) |
| 30-34.9 | −0.306 (−0.614, 0.012) | 0.158 | 0.053 | 0.736 (0.541, 1.012) |
| 35-39.9 | −0.322 (−0.632, <0.001) | 0.160 | 0.044 | 0.725 (0.531, 1.000) |
| 40+ | −0.239 (−0.547, 0.079) | 0.159 | 0.132 | 0.787 (0.579, 1.083) |
| **Charlson Comorbidity Index^a^** | | | | |
| 0 | - | - | - | - |
| 1-2 | −0.066 (−0.192, 0.059) | 0.064 | 0.301 | 0.936 (0.825, 1.061) |
| 3-4 | 0.116 (−0.072, 0.302) | 0.094 | 0.216 | 1.123 (0.931, 1.353) |
| 5+ | 0.116 (−0.129, 0.356) | 0.124 | 0.352 | 1.123 (0.879, 1.427) |
| **Health insurance** | | | | |
| Purchased directly/obtained through employer | - | - | - | - |
| Medicare/Medicaid dual enrollees | −0.038 (−0.309, 0.227) | 0.135 | 0.778 | 0.963 (0.735, 1.254) |
| Medicaid | −0.056 (−0.224, 0.111) | 0.085 | 0.507 | 0.945 (0.799, 1.117) |
| Medicare | −0.172 (−0.382, 0.035) | 0.106 | 0.106 | 0.842 (0.683, 1.035) |
| None | −0.031 (−0.360, 0.286) | 0.163 | 0.847 | 0.969 (0.698, 1.331) |
| Other | −0.027 (−0.308, 0.245) | 0.142 | 0.849 | 0.973 (0.735, 1.277) |
| Skip/prefer not to answer | 0.032 (−0.150, 0.212) | 0.092 | 0.726 | 1.033 (0.861, 1.236) |
| **Highest level of education** | | | | |
| Advanced degree | −0.500 (−0.683, −0.320) | 0.093 | <0.001 | 0.606 (0.505, 0.726) |
| College graduate | −0.210 (−0.364, −0.057) | 0.078 | 0.007 | 0.810 (0.695, 0.944) |
| Some college | - | - | - | - |
| Less than high school | 1.044 (0.844, 1.244) | 0.100 | <0.001 | 2.841 (2.326, 3.469) |
| Prefer not to answer | 0.042 (−0.465, 0.509) | 0.245 | 0.865 | 1.043 (0.628, 1.664) |
| High school or equivalent | 0.417 (0.265, 0.568) | 0.077 | <0.001 | 1.518 (1.304, 1.765) |
| **Household income** | | | | |
| <$50 000 | - | - | - | - |
| $50 000-$100 000 | −0.093 (−0.250, 0.064) | 0.080 | 0.245 | 0.911 (0.779, 1.066) |
| $100 000+ | −0.169 (−0.349, 0.009) | 0.091 | 0.064 | 0.844 (0.706, 1.009) |
| Prefer not to answer/skipped | −0.088 (−0.261, 0.083) | 0.087 | 0.312 | 0.916 (0.771, 1.086) |
| **Health history** | | | | |
| Current or former tobacco use | 0.338 (0.211, 0.467) | 0.065 | <0.001 | 1.402 (1.235, 1.595) |
| History of substance use | 0.525 (0.402, 0.650) | 0.063 | <0.001 | 1.691 (1.495, 1.916) |
| Depression | 0.132 (0.014, 0.249) | 0.060 | 0.027 | 1.141 (1.014, 1.283) |
| Liver disease | 0.003 (−0.135, 0.139) | 0.070 | 0.964 | 1.003 (0.874, 1.150) |
| Hypertension | −0.149 (−0.304, 0.007) | 0.079 | 0.059 | 0.861 (0.738, 1.007) |
| Type 1 diabetes | −0.071 (−0.281, 0.137) | 0.106 | 0.506 | 0.932 (0.755, 1.147) |
| Type 2 diabetes | −0.147 (−0.280, −0.013) | 0.068 | 0.032 | 0.864 (0.756, 0.988) |
| Any off-label non-GLP-1RA AUD medication^b^ | −0.152 (−0.285, −0.020) | 0.067 | 0.024 | 0.859 (0.752, 0.980) |
| Any non-GLP-1RA obesity medication^c^ | 0.022 (−0.092, 0.136) | 0.058 | 0.701 | 1.023 (0.913, 1.146) |
| **Race/ethnicity** | | | | |
| Non-Hispanic (NH) White | - | - | - | - |
| NH Asian | 0.033 (−0.436, 0.471) | 0.234 | 0.886 | 1.034 (0.647, 1.602) |
| NH Black or African American | −0.119 (−0.275, 0.035) | 0.078 | 0.128 | 0.888 (0.760, 1.035) |
| Hispanic | 0.316 (0.160, 0.471) | 0.079 | <0.001 | 1.372 (1.174, 1.602) |
| NH more than one population | −0.118 (−0.377, 0.132) | 0.128 | 0.359 | 0.889 (0.686, 1.141) |
| NH other/none/unknown | −0.244 (−0.590, 0.084) | 0.170 | 0.153 | 0.784 (0.555, 1.087) |
| **Sex** | | | | |
| Female | - | - | - | - |
| Male | 0.681 (0.563, 0.798) | 0.059 | <0.001 | 1.976 (1.757, 2.222) |
| Unknown/other | 0.483 (−0.149, 1.052) | 0.297 | 0.104 | 1.621 (0.862, 2.864) |
| **Visit days** | | | | |
| 1-2 | −0.035 (−0.266, 0.191) | 0.116 | 0.763 | 0.966 (0.766, 1.211) |
| 3-5 | 0.224 (−0.008, 0.452) | 0.117 | 0.054 | 1.252 (0.992, 1.572) |
| 6-10 | 0.268 (0.081, 0.453) | 0.094 | 0.004 | 1.307 (1.084, 1.573) |
| 11-30 | 0.078 (−0.051, 0.207) | 0.066 | 0.236 | 1.081 (0.950, 1.230) |
| 31+ | - | - | - | - |

Abbreviations: AUD, alcohol use disorder; CI, confidence interval; GLP-1RA, glucagon-like peptide-1 receptor agonist; IRR, incidence rate ratio.

^a^ Excludes liver disease and type 2 diabetes.

^b^ Phentermine, phendimetrazine, diethylpropion, phentermine-topiramate, naltrexone-bupropion, orlistat, metformin, topiramate, bupropion.

^c^ Topiramate, gabapentin, baclofen, varenicline, prazosin, doxazosin.

### **eTable 14. Negative Binomial Model Evaluating Association of Alcohol Use with Active GLP-1RA Prescriptions Compared to Future GLP-1RA Prescriptions: AUDIT-C Binge Drinking Question Analysis**

| **Covariate** | **Coefficient (95% CI)** | **Standard error** | ***P* value** | **IRR (95% CI)** |
| --- | --- | --- | --- | --- |
| Active GLP-1RA prescription | −0.038 (−0.111, 0.035) | 0.037 | 0.310 | 0.963 (0.895, 1.035) |
| **Age** | | | | |
| <40 | 0.566 (0.440, 0.692) | 0.064 | <0.001 | 1.761 (1.553, 1.998) |
| 40-49 | 0.375 (0.260, 0.491) | 0.059 | <0.001 | 1.456 (1.297, 1.635) |
| 50-59 | 0.215 (0.109, 0.322) | 0.054 | <0.001 | 1.240 (1.115, 1.380) |
| 60-69 | - | - | - | - |
| 70-79 | −0.408 (−0.591, −0.230) | 0.092 | <0.001 | 0.665 (0.554, 0.795) |
| 80+ | 0.103 (−0.346, 0.504) | 0.216 | 0.633 | 1.108 (0.708, 1.656) |
| **Body mass index** | | | | |
| 18.5-24.9 | - | - | - | - |
| 25-29.9 | 0.044 (−0.180, 0.279) | 0.117 | 0.706 | 1.045 (0.835, 1.321) |
| 30-34.9 | −0.104 (−0.323, 0.125) | 0.114 | 0.360 | 0.901 (0.724, 1.133) |
| 35-39.9 | −0.072 (−0.292, 0.159) | 0.115 | 0.529 | 0.930 (0.747, 1.172) |
| 40+ | −0.085 (−0.304, 0.145) | 0.114 | 0.454 | 0.918 (0.738, 1.156) |
| **Charlson Comorbidity Index^a^** | | | | |
| 0 | - | - | - | - |
| 1-2 | −0.075 (−0.158, 0.008) | 0.042 | 0.078 | 0.928 (0.854, 1.008) |
| 3-4 | −0.076 (−0.204, 0.050) | 0.065 | 0.240 | 0.927 (0.815, 1.052) |
| 5+ | −0.037 (−0.205, 0.126) | 0.084 | 0.660 | 0.963 (0.815, 1.135) |
| **Health insurance** | | | | |
| Purchased directly/obtained through employer | - | - | - | - |
| Medicare/Medicaid dual enrollees | −0.342 (−0.540, −0.150) | 0.099 | 0.001 | 0.711 (0.583, 0.861) |
| Medicaid | −0.127 (−0.240, −0.014) | 0.057 | 0.027 | 0.881 (0.787, 0.986) |
| Medicare | −0.299 (−0.441, −0.160) | 0.072 | <0.001 | 0.741 (0.644, 0.852) |
| None | 0.020 (−0.194, 0.226) | 0.107 | 0.850 | 1.020 (0.824, 1.253) |
| Other | −0.113 (−0.307, 0.074) | 0.097 | 0.246 | 0.893 (0.736, 1.077) |
| Skip/prefer not to answer | 0.008 (−0.113, 0.127) | 0.061 | 0.895 | 1.008 (0.894, 1.135) |
| **Highest level of education** | | | | |
| Advanced degree | −0.268 (−0.384, −0.153) | 0.059 | <0.001 | 0.765 (0.681, 0.859) |
| College graduate | −0.166 (−0.269, −0.063) | 0.052 | 0.002 | 0.847 (0.764, 0.939) |
| Some college | - | - | - | - |
| Less than high school | 0.669 (0.530, 0.805) | 0.070 | <0.001 | 1.951 (1.699, 2.237) |
| Prefer not to answer | −0.050 (−0.384, 0.257) | 0.163 | 0.757 | 0.951 (0.681, 1.293) |
| High school or equivalent | 0.344 (0.241, 0.446) | 0.052 | <0.001 | 1.410 (1.273, 1.561) |
| **Household income** | | | | |
| <$50 000 | - | - | - | - |
| $50 000-$100 000 | −0.069 (−0.175, 0.036) | 0.054 | 0.199 | 0.933 (0.840, 1.037) |
| $100 000+ | 0.006 (−0.112, 0.124) | 0.060 | 0.919 | 1.006 (0.894, 1.131) |
| Prefer not to answer/skipped | −0.023 (−0.138, 0.090) | 0.058 | 0.690 | 0.977 (0.871, 1.094) |
| **Health history** | | | | |
| Current or former tobacco use | 0.243 (0.159, 0.327) | 0.043 | <0.001 | 1.275 (1.172, 1.387) |
| History of substance use | 0.292 (0.211, 0.374) | 0.041 | <0.001 | 1.340 (1.236, 1.453) |
| Depression | 0.125 (0.047, 0.202) | 0.040 | 0.002 | 1.133 (1.048, 1.224) |
| Liver disease | 0.099 (0.007, 0.189) | 0.046 | 0.033 | 1.104 (1.007, 1.209) |
| Hypertension | −0.072 (−0.176, 0.033) | 0.053 | 0.176 | 0.930 (0.839, 1.033) |
| Type 1 diabetes | −0.166 (−0.308, −0.026) | 0.072 | 0.021 | 0.847 (0.735, 0.974) |
| Type 2 diabetes | −0.170 (−0.259, −0.081) | 0.045 | <0.001 | 0.844 (0.772, 0.922) |
| Any off-label non-GLP-1RA AUD medication^b^ | 0.050 (−0.037, 0.136) | 0.044 | 0.260 | 1.051 (0.963, 1.146) |
| Any non-GLP-1RA obesity medication^c^ | −0.031 (−0.107, 0.045) | 0.039 | 0.420 | 0.969 (0.898, 1.046) |
| **Race/ethnicity** | | | | |
| Non-Hispanic (NH) White | - | - | - | - |
| NH Asian | −0.282 (−0.650, 0.051) | 0.179 | 0.114 | 0.754 (0.522, 1.053) |
| NH Black or African American | 0.212 (0.114, 0.310) | 0.050 | <0.001 | 1.237 (1.121, 1.364) |
| Hispanic | 0.216 (0.107, 0.324) | 0.055 | <0.001 | 1.242 (1.113, 1.383) |
| NH more than one population | −0.041 (−0.215, 0.127) | 0.087 | 0.639 | 0.960 (0.806, 1.135) |
| NH other/none/unknown | 0.177 (−0.027, 0.373) | 0.102 | 0.081 | 1.194 (0.973, 1.452) |
| **Sex** | | | | |
| Female | - | - | - | - |
| Male | 0.437 (0.358, 0.515) | 0.040 | <0.001 | 1.547 (1.430, 1.674) |
| Unknown/Other | 0.561 (0.207, 0.886) | 0.174 | 0.001 | 1.752 (1.230, 2.426) |
| **Visit days** | | | | |
| 1-2 | −0.031 (−0.185, 0.119) | 0.078 | 0.690 | 0.970 (0.831, 1.127) |
| 3-5 | 0.142 (−0.016, 0.295) | 0.079 | 0.075 | 1.152 (0.984, 1.344) |
| 6-10 | 0.133 (0.006, 0.258) | 0.064 | 0.038 | 1.142 (1.006, 1.294) |
| 11-30 | 0.080 (−0.006, 0.165) | 0.044 | 0.069 | 1.083 (0.994, 1.180) |
| 31+ | - | - | - | - |

Abbreviations: AUD, alcohol use disorder; CI, confidence interval; GLP-1RA, glucagon-like peptide-1 receptor agonist; IRR, incidence rate ratio.

^a^ Excludes liver disease and type 2 diabetes.

^b^ Phentermine, phendimetrazine, diethylpropion, phentermine-topiramate, naltrexone-bupropion, orlistat, metformin, topiramate, bupropion.

^c^ Topiramate, gabapentin, baclofen, varenicline, prazosin, doxazosin.

### **eTable 15. Negative Binomial Model Evaluating Association of Alcohol Use with Former GLP-1RA Prescriptions Compared to Future GLP-1RA Prescriptions: AUDIT-C Drink Frequency Question Analysis**

| **Covariate** | **Coefficient (95% CI)** | **Standard error** | ***P* value** | **IRR (95% CI)** |
| --- | --- | --- | --- | --- |
| Former GLP-1RA prescription | −0.109 (−0.195, −0.026) | 0.043 | 0.011 | 0.897 (0.823, 0.975) |
| **Age** | | | | |
| <40 | 0.112 (0.032, 0.191) | 0.040 | 0.006 | 1.118 (1.033, 1.211) |
| 40-49 | 0.037 (−0.036, 0.110) | 0.037 | 0.323 | 1.037 (0.964, 1.116) |
| 50-59 | 0.050 (−0.013, 0.115) | 0.033 | 0.122 | 1.052 (0.987, 1.121) |
| 60-69 | - | - | - | - |
| 70-79 | 0.049 (−0.036, 0.133) | 0.043 | 0.256 | 1.050 (0.965, 1.142) |
| 80+ | 0.066 (−0.174, 0.292) | 0.119 | 0.575 | 1.069 (0.840, 1.339) |
| **Body mass index** | | | | |
| 18.5-24.9 | - | - | - | - |
| 25-29.9 | −0.100 (−0.228, 0.031) | 0.066 | 0.130 | 0.905 (0.796, 1.032) |
| 30-34.9 | −0.143 (−0.266, −0.016) | 0.064 | 0.025 | 0.867 (0.766, 0.985) |
| 35-39.9 | −0.162 (−0.286, −0.033) | 0.065 | 0.012 | 0.851 (0.751, 0.968) |
| 40+ | −0.270 (−0.396, −0.141) | 0.065 | <0.001 | 0.763 (0.673, 0.868) |
| **Charlson Comorbidity Index^a^** | | | | |
| 0 | - | - | - | - |
| 1-2 | 0.026 (−0.026, 0.078) | 0.026 | 0.320 | 1.027 (0.975, 1.081) |
| 3-4 | 0.043 (−0.033, 0.119) | 0.039 | 0.267 | 1.044 (0.967, 1.126) |
| 5+ | −0.085 (−0.190, 0.018) | 0.053 | 0.107 | 0.918 (0.827, 1.018) |
| **Health insurance** | | | | |
| Purchased directly/obtained through employer | - | - | - | - |
| Medicare/Medicaid dual enrollees | −0.280 (−0.410, −0.154) | 0.065 | <0.001 | 0.756 (0.664, 0.858) |
| Medicaid | −0.081 (−0.157, −0.005) | 0.039 | 0.036 | 0.922 (0.855, 0.995) |
| Medicare | −0.024 (−0.101, 0.053) | 0.039 | 0.548 | 0.977 (0.904, 1.055) |
| None | −0.058 (−0.217, 0.095) | 0.080 | 0.467 | 0.944 (0.805, 1.100) |
| Other/prefer not to answer | −0.066 (−0.133, 0.001) | 0.034 | 0.053 | 0.936 (0.875, 1.001) |
| **Highest level of education** | | | | |
| Advanced degree | 0.128 (0.063, 0.193) | 0.033 | <0.001 | 1.137 (1.065, 1.213) |
| College graduate | 0.064 (0.002, 0.125) | 0.031 | 0.043 | 1.066 (1.002, 1.133) |
| Some college | - | - | - | - |
| Less than high school | −0.055 (−0.167, 0.053) | 0.056 | 0.323 | 0.946 (0.847, 1.055) |
| Prefer not to answer | −0.045 (−0.249, 0.148) | 0.101 | 0.656 | 0.956 (0.779, 1.160) |
| High school or equivalent | −0.026 (−0.098, 0.045) | 0.037 | 0.477 | 0.974 (0.907, 1.046) |
| **Household income** |  |  |  |  |
| <$50 000 | - | - | - | - |
| $50 000-$100 000 | 0.127 (0.061, 0.193) | 0.034 | <0.001 | 1.135 (1.063, 1.213) |
| $100 000+ | 0.277 (0.206, 0.348) | 0.036 | <0.001 | 1.319 (1.228, 1.416) |
| Prefer not to answer/skipped | 0.040 (−0.036, 0.116) | 0.039 | 0.299 | 1.041 (0.964, 1.123) |
| **Health history** | | | | |
| Current or former tobacco use | 0.119 (0.069, 0.169) | 0.026 | <0.001 | 1.126 (1.072, 1.184) |
| History of substance use | 0.168 (0.119, 0.218) | 0.025 | <0.001 | 1.183 (1.127, 1.243) |
| Depression | −0.048 (−0.096, 0.001) | 0.025 | 0.055 | 0.954 (0.908, 1.001) |
| Liver disease | −0.005 (−0.066, 0.055) | 0.031 | 0.874 | 0.995 (0.936, 1.057) |
| Hypertension | 0.026 (−0.040, 0.092) | 0.034 | 0.444 | 1.026 (0.961, 1.097) |
| Type 1 diabetes | −0.213 (−0.309, −0.119) | 0.048 | <0.001 | 0.808 (0.734, 0.888) |
| Type 2 diabetes | −0.156 (−0.210, −0.102) | 0.027 | <0.001 | 0.855 (0.811, 0.903) |
| Any off-label non-GLP-1RA AUD medication^b^ | −0.070 (−0.127, −0.014) | 0.029 | 0.015 | 0.932 (0.881, 0.986) |
| Any non-GLP-1RA obesity medication^c^ | −0.023 (−0.071, 0.025) | 0.025 | 0.348 | 0.977 (0.931, 1.025) |
| **Race/ethnicity** | | | | |
| Non-Hispanic (NH) White | - | - | - | - |
| NH Asian | −0.217 (−0.418, −0.027) | 0.100 | 0.030 | 0.805 (0.658, 0.974) |
| NH Black or African American | 0.032 (−0.031, 0.094) | 0.032 | 0.322 | 1.032 (0.969, 1.099) |
| Hispanic | −0.107 (−0.185, −0.030) | 0.039 | 0.007 | 0.898 (0.831, 0.970) |
| NH more than one population | −0.027 (−0.138, 0.080) | 0.056 | 0.627 | 0.973 (0.871, 1.084) |
| NH other/none/unknown | −0.181 (−0.318, −0.048) | 0.069 | 0.009 | 0.835 (0.727, 0.953) |
| **Sex** | | | | |
| Female | - | - | - | - |
| Male | 0.094 (0.044, 0.145) | 0.026 | <0.001 | 1.099 (1.045, 1.156) |
| Unknown/other | 0.271 (0.030, 0.494) | 0.118 | 0.022 | 1.311 (1.031, 1.639) |
| **Visit days** | | | | |
| 1-2 | −0.070 (−0.168, 0.026) | 0.049 | 0.155 | 0.932 (0.845, 1.026) |
| 3-5 | 0.065 (−0.030, 0.158) | 0.048 | 0.177 | 1.067 (0.970, 1.171) |
| 6-10 | 0.041 (−0.037, 0.118) | 0.040 | 0.300 | 1.042 (0.964, 1.125) |
| 11-30 | 0.067 (0.013, 0.120) | 0.027 | 0.014 | 1.069 (1.014, 1.127) |
| 31+ | - | - | - | - |

Abbreviations: AUD, alcohol use disorder; CI, confidence interval; GLP-1RA, glucagon-like peptide-1 receptor agonist; IRR, incidence rate ratio.

^a^ Excludes liver disease and type 2 diabetes.

^b^ Phentermine, phendimetrazine, diethylpropion, phentermine-topiramate, naltrexone-bupropion, orlistat, metformin, topiramate, bupropion.

^c^ Topiramate, gabapentin, baclofen, varenicline, prazosin, doxazosin.

### **eTable 16. Negative Binomial Model Evaluating Association of Alcohol Use with Former GLP-1RA Prescriptions Compared to Future GLP-1RA Prescriptions: AUDIT-C Drink Quantity Question Analysis**

| **Covariate** | **Coefficient (95% CI)** | **Standard error** | ***P* value** | **IRR (95% CI)** |
| --- | --- | --- | --- | --- |
| Former GLP-1RA prescription | 0.019 (−0.212, 0.242) | 0.114 | 0.872 | 1.019 (0.809, 1.274) |
| **Age** | | | | |
| <40 | 0.790 (0.571, 1.011) | 0.112 | <0.001 | 2.204 (1.770, 2.749) |
| 40-49 | 0.357 (0.144, 0.572) | 0.109 | 0.001 | 1.430 (1.155, 1.771) |
| 50-59 | 0.448 (0.258, 0.640) | 0.098 | <0.001 | 1.565 (1.294, 1.897) |
| 60-69 | - | - | - | - |
| 70-79 | −0.727 (−1.079, −0.392) | 0.177 | <0.001 | 0.483 (0.340, 0.676) |
| 80+ | −1.177 (−2.540, −0.154) | 0.620 | 0.058 | 0.308 (0.079, 0.858) |
| **Body mass index** | | | | |
| 18.5-24.9 | **-** | **-** | **-** | **-** |
| 25-29.9 | −0.227 (−0.612, 0.170) | 0.196 | 0.246 | 0.797 (0.542, 1.185) |
| 30-34.9 | −0.227 (−0.595, 0.156) | 0.188 | 0.227 | 0.797 (0.551, 1.168) |
| 35-39.9 | −0.270 (−0.642, 0.116) | 0.190 | 0.155 | 0.763 (0.526, 1.123) |
| 40+ | −0.255 (−0.625, 0.128) | 0.189 | 0.176 | 0.775 (0.535, 1.136) |
| **Charlson Comorbidity Index^a^** | | | | |
| 0 | - | - | - | - |
| 1-2 | −0.048 (−0.198, 0.102) | 0.076 | 0.529 | 0.953 (0.821, 1.107) |
| 3-4 | 0.068 (−0.157, 0.289) | 0.111 | 0.544 | 1.070 (0.855, 1.335) |
| 5+ | 0.231 (−0.057, 0.512) | 0.145 | 0.113 | 1.259 (0.945, 1.669) |
| **Health insurance** | | | | |
| Purchased directly/obtained through employer | - | - | - | - |
| Medicare/Medicaid dual enrollees | 0.106 (−0.221, 0.424) | 0.163 | 0.517 | 1.112 (0.802, 1.528) |
| Medicaid | −0.077 (−0.275, 0.120) | 0.100 | 0.440 | 0.926 (0.760, 1.127) |
| Medicare | −0.124 (−0.371, 0.119) | 0.126 | 0.323 | 0.883 (0.690, 1.127) |
| None | −0.119 (−0.497, 0.245) | 0.186 | 0.523 | 0.888 (0.609, 1.278) |
| Other/prefer not to answer | −0.050 (−0.240, 0.137) | 0.096 | 0.602 | 0.951 (0.787, 1.147) |
| **Highest level of education** | | | | |
| Advanced degree | −0.484 (−0.707, −0.265) | 0.113 | <0.001 | 0.616 (0.493, 0.767) |
| College graduate | −0.234 (−0.419, −0.050) | 0.094 | 0.013 | 0.791 (0.657, 0.951) |
| Some college | - | - | - | - |
| Less than high school | 0.834 (0.595, 1.072) | 0.120 | <0.001 | 2.303 (1.814, 2.920) |
| Prefer not to answer | 0.212 (−0.333, 0.721) | 0.260 | 0.415 | 1.236 (0.716, 2.058) |
| High school or equivalent | 0.410 (0.231, 0.588) | 0.090 | <0.001 | 1.507 (1.260, 1.801) |
| **Household income** | | | | |
| <$50 000 | - | - | - | - |
| $50 000-$100 000 | −0.125 (−0.315, 0.064) | 0.096 | 0.195 | 0.883 (0.730, 1.066) |
| $100 000+ | −0.192 (−0.407, 0.022) | 0.110 | 0.080 | 0.825 (0.665, 1.022) |
| Prefer not to answer/skipped | −0.112 (−0.318, 0.091) | 0.104 | 0.280 | 0.894 (0.727, 1.095) |
| **Health history** |  |  |  |  |
| Current or former tobacco use | 0.481 (0.330, 0.634) | 0.078 | <0.001 | 1.618 (1.392, 1.885) |
| History of substance use | 0.405 (0.259, 0.552) | 0.075 | <0.001 | 1.499 (1.295, 1.737) |
| Depression | 0.092 (−0.050, 0.234) | 0.072 | 0.203 | 1.096 (0.951, 1.263) |
| Liver disease | −0.063 (−0.239, 0.111) | 0.089 | 0.483 | 0.939 (0.787, 1.118) |
| Hypertension | −0.061 (−0.243, 0.124) | 0.093 | 0.513 | 0.941 (0.784, 1.132) |
| Type 1 diabetes | −0.129 (−0.401, 0.137) | 0.136 | 0.345 | 0.879 (0.670, 1.147) |
| Type 2 diabetes | −0.120 (−0.275, 0.036) | 0.079 | 0.127 | 0.887 (0.759, 1.036) |
| Any off-label non-GLP-1RA AUD medication^b^ | −0.183 (−0.348, −0.020) | 0.084 | 0.029 | 0.833 (0.706, 0.981) |
| Any non-GLP-1RA obesity medication^c^ | 0.020 (−0.119, 0.158) | 0.070 | 0.780 | 1.020 (0.888, 1.171) |
| **Race/ethnicity** | | | | |
| Non-Hispanic (NH) White | - | - | - | - |
| NH Asian | 0.072 (−0.529, 0.623) | 0.297 | 0.808 | 1.075 (0.589, 1.864) |
| NH Black or African American | −0.092 (−0.273, 0.087) | 0.091 | 0.312 | 0.912 (0.761, 1.090) |
| Hispanic | 0.310 (0.120, 0.498) | 0.095 | 0.001 | 1.363 (1.127, 1.646) |
| NH more than one population | 0.084 (−0.216, 0.373) | 0.148 | 0.570 | 1.087 (0.806, 1.452) |
| NH other/none/unknown | −0.194 (−0.602, 0.191) | 0.203 | 0.339 | 0.823 (0.548, 1.210) |
| **Sex** | | | | |
| Female | - | - | - | - |
| Male | 0.642 (0.502, 0.782) | 0.070 | <0.001 | 1.900 (1.652, 2.185) |
| Unknown/other | 0.500 (−0.264, 1.181) | 0.367 | 0.173 | 1.649 (0.768, 3.258) |
| **Visit days** | | | | |
| 1-2 | 0.107 (−0.162, 0.372) | 0.135 | 0.425 | 1.113 (0.850, 1.450) |
| 3-5 | 0.126 (−0.140, 0.386) | 0.133 | 0.346 | 1.134 (0.869, 1.471) |
| 6-10 | 0.152 (−0.071, 0.373) | 0.112 | 0.175 | 1.165 (0.931, 1.452) |
| 11-30 | 0.041 (−0.114, 0.196) | 0.079 | 0.603 | 1.042 (0.892, 1.216) |
| 31+ | - | - | - | - |

Abbreviations: AUD, alcohol use disorder; CI, confidence interval; GLP-1RA, glucagon-like peptide-1 receptor agonist; IRR, incidence rate ratio.

^a^ Excludes liver disease and type 2 diabetes.

^b^ Phentermine, phendimetrazine, diethylpropion, phentermine-topiramate, naltrexone-bupropion, orlistat, metformin, topiramate, bupropion.

^c^ Topiramate, gabapentin, baclofen, varenicline, prazosin, doxazosin.

### **eTable 17. Negative Binomial Model Evaluating Association of Alcohol Use with Former GLP-1RA Prescriptions Compared to Future GLP-1RA Prescriptions: AUDIT-C Binge Drinking Question Analysis**

| **Covariate** | **Coefficient (95% CI)** | **Standard error** | ***P* value** | **IRR (95% CI)** |
| --- | --- | --- | --- | --- |
| Former GLP-1RA prescription | 0.043 (−0.107, 0.189) | 0.076 | 0.566 | 1.044 (0.898, 1.208) |
| **Age** | | | | |
| <40 | 0.618 (0.471, 0.766) | 0.075 | <0.001 | 1.855 (1.601, 2.151) |
| 40-49 | 0.383 (0.243, 0.524) | 0.072 | <0.001 | 1.467 (1.276, 1.689) |
| 50-59 | 0.280 (0.151, 0.410) | 0.066 | <0.001 | 1.323 (1.163, 1.506) |
| 60-69 | - | - | - | - |
| 70-79 | −0.385 (−0.610, −0.166) | 0.113 | 0.001 | 0.681 (0.543, 0.847) |
| 80+ | 0.223 (−0.317, 0.697) | 0.256 | 0.385 | 1.249 (0.728, 2.008) |
| **Body mass index** | | | | |
| 18.5-24.9 | - | - | - | - |
| 25-29.9 | 0.032 (−0.227, 0.304) | 0.135 | 0.814 | 1.032 (0.797, 1.356) |
| 30-34.9 | −0.059 (−0.309, 0.205) | 0.131 | 0.650 | 0.942 (0.734, 1.228) |
| 35-39.9 | −0.113 (−0.366, 0.154) | 0.132 | 0.393 | 0.893 (0.694, 1.167) |
| 40+ | −0.172 (−0.424, 0.094) | 0.132 | 0.191 | 0.842 (0.654, 1.098) |
| **Charlson Comorbidity Index^a^** | | | | |
| 0 | - | - | - | - |
| 1-2 | −0.021 (−0.120, 0.078) | 0.050 | 0.675 | 0.979 (0.887, 1.081) |
| 3-4 | −0.048 (−0.199, 0.100) | 0.076 | 0.528 | 0.953 (0.820, 1.105) |
| 5+ | 0.064 (−0.133, 0.256) | 0.099 | 0.517 | 1.066 (0.875, 1.292) |
| **Health insurance** | | | | |
| Purchased directly/obtained through employer | - | - | - | - |
| Medicare/Medicaid dual enrollees | −0.306 (−0.554, −0.069) | 0.123 | 0.013 | 0.736 (0.575, 0.933) |
| Medicaid | −0.128 (−0.260, 0.003) | 0.067 | 0.056 | 0.880 (0.771, 1.003) |
| Medicare | −0.257 (−0.425, −0.091) | 0.085 | 0.003 | 0.774 (0.654, 0.913) |
| None | −0.103 (−0.354, 0.137) | 0.125 | 0.410 | 0.902 (0.702, 1.146) |
| Other/prefer not to answer | −0.066 (−0.192, 0.058) | 0.064 | 0.297 | 0.936 (0.826, 1.059) |
| **Highest level of education** | | | | |
| Advanced degree | −0.193 (−0.335, −0.053) | 0.072 | 0.007 | 0.824 (0.715, 0.948) |
| College graduate | −0.111 (−0.235, 0.012) | 0.063 | 0.078 | 0.895 (0.791, 1.012) |
| Some college | - | - | - | - |
| Less than high school | 0.604 (0.441, 0.766) | 0.083 | <0.001 | 1.830 (1.554, 2.151) |
| Prefer not to answer | 0.188 (−0.165, 0.515) | 0.172 | 0.274 | 1.207 (0.848, 1.674) |
| High school or equivalent | 0.390 (0.269, 0.511) | 0.061 | <0.001 | 1.478 (1.309, 1.666) |
| **Household income** | | | | |
| <$50 000 | - | - | - | - |
| $50 000-$100 000 | −0.105 (−0.233, 0.022) | 0.065 | 0.105 | 0.900 (0.792, 1.022) |
| $100 000+ | −0.041 (−0.183, 0.101) | 0.072 | 0.572 | 0.960 (0.833, 1.106) |
| Prefer not to answer/skipped | −0.064 (−0.201, 0.070) | 0.069 | 0.353 | 0.938 (0.818, 1.073) |
| **Health history** | | | | |
| Current or former tobacco use | 0.255 (0.157, 0.354) | 0.050 | <0.001 | 1.291 (1.170, 1.425) |
| History of substance use | 0.252 (0.156, 0.348) | 0.049 | <0.001 | 1.286 (1.169, 1.416) |
| Depression | 0.094 (<0.001, 0.187) | 0.048 | 0.049 | 1.099 (1.000, 1.206) |
| Liver disease | 0.038 (−0.078, 0.152) | 0.059 | 0.521 | 1.038 (0.925, 1.164) |
| Hypertension | −0.015 (−0.136, 0.109) | 0.063 | 0.815 | 0.985 (0.872, 1.115) |
| Type 1 diabetes | −0.101 (−0.280, 0.073) | 0.090 | 0.261 | 0.904 (0.756, 1.076) |
| Type 2 diabetes | −0.123 (−0.226, −0.020) | 0.052 | 0.019 | 0.884 (0.798, 0.980) |
| Any off-label non-GLP-1RA AUD medication^b^ | 0.025 (−0.082, 0.131) | 0.054 | 0.641 | 1.026 (0.922, 1.140) |
| Any non-GLP-1RA obesity medication^c^ | −0.012 (−0.103, 0.080) | 0.047 | 0.805 | 0.989 (0.902, 1.084) |
| **Race/ethnicity** | | | | |
| Non-Hispanic (NH) White | - | - | - | - |
| NH Asian | −0.480 (−0.998, −0.030) | 0.246 | 0.051 | 0.619 (0.369, 0.970) |
| NH Black or African American | 0.217 (0.102, 0.332) | 0.059 | <0.001 | 1.243 (1.107, 1.394) |
| Hispanic | 0.265 (0.135, 0.393) | 0.066 | <0.001 | 1.303 (1.145, 1.482) |
| NH more than one population | −0.023 (−0.237, 0.181) | 0.107 | 0.830 | 0.977 (0.789, 1.199) |
| NH other/none/unknown | 0.230 (−0.014, 0.462) | 0.121 | 0.057 | 1.259 (0.986, 1.587) |
| **Sex** | | | | |
| Female | - | - | - | - |
| Male | 0.419 (0.325, 0.513) | 0.048 | <0.001 | 1.521 (1.385, 1.670) |
| Unknown/other | 0.616 (0.182, 1.009) | 0.211 | 0.003 | 1.852 (1.199, 2.744) |
| **Visit days** | | | | |
| 1-2 | 0.050 (−0.132, 0.227) | 0.092 | 0.588 | 1.051 (0.876, 1.254) |
| 3-5 | 0.043 (−0.139, 0.220) | 0.092 | 0.642 | 1.043 (0.870, 1.246) |
| 6-10 | 0.008 (−0.145, 0.158) | 0.077 | 0.914 | 1.008 (0.865, 1.172) |
| 11-30 | 0.072 (−0.030, 0.173) | 0.052 | 0.167 | 1.074 (0.970, 1.189) |
| 31+ | - | - | - | - |

Abbreviations: AUD, alcohol use disorder; CI, confidence interval; GLP-1RA, glucagon-like peptide-1 receptor agonist; IRR, incidence rate ratio.

^a^ Excludes liver disease and type 2 diabetes.

^b^ Phentermine, phendimetrazine, diethylpropion, phentermine-topiramate, naltrexone-bupropion, orlistat, metformin, topiramate, bupropion.

^c^ Topiramate, gabapentin, baclofen, varenicline, prazosin, doxazosin.

### **eTable 18. GLP-1RA Types for the Active, Former and Future Prescription Groups**

| **GLP-1RA Type** | **N (%) Active GLP-1RA** | **N (%) Former GLP-1RA** | **N (%) Future GLP-1RA** |
| --- | --- | --- | --- |
| Albiglutide | 59 (1.6%) | <20 | <20 |
| Dulaglutide | 1799 (49.3%) | 231 (42.5%) | 2175 (38.6%) |
| Exenatide | 465 (12.7%) | 65 (11.9%) | 152 (2.7%) |
| Liraglutide | 1595 (43.7%) | 187 (34.4%) | 1052 (18.6%) |
| Lixisenatide | 25 (0.7%) | <20 | <20 |
| Semaglutide | 2074 (56.8%) | 257 (47.2%) | 3412 (60.5%) |
| Tirzepatide | 393 (10.8%) | <20 | 524 (9.3%) |
